# Machine Learning Analysis of Post-Acute COVID Symptoms Identifies Distinct Clusters, Severity Groups, and Trajectories

**DOI:** 10.1101/2025.11.16.25340350

**Authors:** Beverly Peng, Thomas Dalhuisen, Aidan Rogers, Violeta Capric, Helen E. Davies, Steven G. Deeks, Christopher L. Dupont, Jesus Estevez, Marcelo Freire, Natalie S. Haddad, Samantha A. Jones, J. Daniel Kelly, Kristin Ladell, F. Eun-Hyung Lee, Jeffrey N. Martin, Kelly L. Miners, Michael J. Peluso, David A. Price, Amy D. Proal, David Putrino, Richard H. Scheuermann, Michael B. VanElzakker, Yun Zhang, Gene S. Tan, Yu Qian

**Affiliations:** J. Craig Venter Institute, La Jolla, CA, USA; Division of HIV, Infectious Diseases, and Global Medicine, University of California, San Francisco, San Francisco, CA, USA; Department of Rehabilitation and Human Performance, Icahn School of Medicine at Mount Sinai, New York, NY, USA; Division of Pulmonary, Allergy, Critical Care, and Sleep Medicine, Department of Medicine, Emory University, Atlanta, GA, USA; Department of Respiratory Medicine, University Hospital of Wales, Cardiff, UK; Department of Dermatology, School of Medicine, University of California, San Diego, La Jolla, CA, USA; Division of Infectious Diseases and Global Public Health, Department of Medicine, University of California, San Diego, La Jolla, CA, USA; Department of Epidemiology and Biostatistics, University of California, San Francisco, San Francisco, CA, USA; Institute for Global Health Sciences, University of California, San Francisco, San Francisco, CA, USA; Division of Infection and Immunity, Cardiff University School of Medicine, University Hospital of Wales, Cardiff, UK; Lowance Center for Human Immunology, Emory University, Atlanta, GA, USA; Systems Immunity Research Institute, Cardiff University School of Medicine, University Hospital of Wales, Cardiff, UK; PolyBio Research Foundation, Medford, MA, USA; Division of Intramural Research, National Library of Medicine, National Institutes of Health, Bethesda, MD, USA; Division of Neurotherapeutics, Massachusetts General Hospital, Boston, MA, USA; Herbert Wertheim School of Public Health and Human Longevity Science, University of California, San Diego, La Jolla, CA, USA

## Abstract

Questionnaires that capture patient-reported symptomatology provide low-cost but potentially high-value data for the *de novo* discovery of disease phenotype, severity, and responsiveness to intervention groupings within an umbrella condition. The availability of comprehensive electronic health records (EHRs) has nonetheless overshadowed the use of questionnaires data for symptom analysis in the context of COVID-19. We analyzed de-identified questionnaires from post-acute COVID-19 cohorts at the University of California, San Francisco (UCSF, n = 669), Icahn School of Medicine at Mount Sinai (ISMMS, n = 615), Emory University (Emory, n = 60), and the University Hospital of Wales (Cardiff, n = 317). Using topic modeling followed by unsupervised clustering, we identified distinct symptom clusters and their corresponding symptom signatures. Mapping these signatures to organ systems revealed nine to twelve endotypes per cohort, capturing the heterogeneity of post-COVID-19 symptoms. Some clusters were associated with pre-existing conditions, including a female-predominant severity cluster with neurological and hormonal symptoms. Longitudinal analysis distinguished three symptom trajectories of individual symptoms: acute then resolving, persistent but attenuated, and progressive disease. Symptom severity correlates between the acute and the long COVID phases. Individuals with non-mild symptoms in the acute phase have 2.6 times the risk of developing moderate or severe long COVID symptoms, compared with individuals with mild symptoms in the acute phase. Across all cohorts, three severity levels, namely, mild, moderate, and severe, were evident from symptoms alone. Symptom-based severity scores correlated with patient-reported health status (EQ-5D) and SARS-CoV-2-specific antibody responses in plasmablasts, validating the prediction. Cluster-level analyses further stratified patients into recovered and non-recovered subgroups, identifying endotypes associated with different recovery trajectories. Finally, meta-analysis integrating cohort-specific clusters defined ten global endotypes and a unified map of severity scores, highlighting cohort-specific patterns, sex differences, and relationships among organ systems. These findings demonstrate that machine learning-assisted screening of questionnaire data can robustly identify symptom clusters, endotypes, and severity groups, providing a framework for stratifying long COVID patients for precision medicine trial design.

## 1. Introduction

Severe acute respiratory syndrome coronavirus 2 (SARS-CoV-2) infection can have long-term effects on health. Post-acute sequelae of SARS-CoV-2 infection (PASC), alternatively known as long coronavirus disease (long COVID), is a condition defined by the World Health Organization (WHO) as unexplained symptoms that start within 3 months and last at least 2 months after infection with SARS-CoV-2 [Soriano 2022] and by the National Academies of Science, Engineering, and Medicine as an infection-associated chronic condition (IACC) with a continuous, relapsing and remitting, or progressive disease course that affects one or more organ systems and persists for at least 3 months after infection with SARS-CoV-2 [NASEM 2024]. The wide array of reported symptoms has been a major challenge for studies of long COVID [Lopez-Leon 2021, Davis 2023, Chippa 2024]. Moreover, while some symptoms are resolved within weeks to months, others persist for years. The origins, trajectories, and interrelationships of these heterogeneous symptoms remain poorly understood. Most investigators agree that multiple organ systems and interacting mechanisms underlie long COVID, including dynamic interactions between pre-existing host immunity and possible viral reservoirs in tissues [Constantinescu-Bercu 2023, Peluso 2024a-b, Swank 2024], such as the endemic herpesviridae [Gáspár 2025, Mak 2025] or persistent SARS-CoV-2 [Proal 2023, Proal 2025, Prakash 2025]. Before designing mechanistic studies or clinical trials targeting affected organ systems, it is essential to characterize and categorize the complexity and heterogeneity of post-infection symptom profiles.

Disease phenotyping efforts to date have often relied on the construction of symptom groupings based on clinical intuition [Frontera 2022, Thaweethai 2023, Geng 2024]. Recent advances in artificial intelligence (AI) and machine learning (ML) provide more objective methods to detect and characterize long COVID. For example, supervised classification algorithms including XGBoost and Random Forest have been applied to distinguish patients with long COVID [Ahmad 2024] or identify risk factors (e.g., sex and age groups) [Hill 2023, O’Neil 2024] associated with long COVID from electronic health records (EHRs). Four reproducible long COVID subphenotypes have been reported from over 137 symptoms and conditions described in EHRs [Zhang 2023]. Longitudinal statistical analyses have further shown that hospitalization during the acute phase of SARS-CoV-2 infection is a risk factor for certain long-term effects [Bowe 2023], although it is unclear these effects are due to post-intensive care syndrome or long COVID. These subphenotypes and risk factors can potentially be used to define endotypes and severity groups among patients with long COVID. However, it is still challenging to predict such endotypes and severity groups using AI/ML, partially due to the use of EHRs. Much of the existing literature has used EHRs to focus on long COVID subphenotypes, providing conceptual evidence for organ system-specific subgroups, but there are inherent limitations to this approach, including missing data, ambiguity, varying fields and formats across sites, and time burden.

An alternative and more accessible source of information is standardized symptom questionnaires, which are quick and relatively easy to acquire. These questionnaires typically capture binary responses for approximately 30–60 common symptoms and are widely available across clinical settings, including those with limited technological resources. Unlike the EHRs that usually have highly variable formats, questionnaire data are inherently standardized but relatively limited in size. Computational models built on questionnaire data are generally more interpretable due to their simplicity and can be readily applied to prospective screening through ML-based classification, without requiring specialized medical expertise or access to comprehensive EHR. If questionnaire data prove to be informative, new patients could be rapidly profiled for clinical trial eligibility and/or treatment stratification, even on their first visit. A key limitation is that questionnaire datasets are often small, raising uncertainty about whether robust patterns can be identified consistently.

This study focuses on unsupervised analyses of questionnaire-based symptoms, with three specific aims: (1) to determine whether patients can be grouped by symptom profiles and whether signature symptoms can be identified for each group; (2) to incorporate organ-system relevance into symptom analyses to define biologically meaningful endotypes; and (3) to derive quantitative severity scores from questionnaire symptoms that can be mapped to qualitative severity levels and plasma or serum-based metrics. The first goal creates a computational challenge driven by the curse of dimensionality: when every symptom is used as a separate feature, each patient can appear unique, and clustering becomes unfeasible. Existing dimensionality reduction methods including principal component analysis (PCA) and uniform manifold approximation and projection (UMAP) are designed for data visualization, which present most variances in the input data using only two or three latent variables. Recent work has shown that PCA does not help identify symptom clusters [Fresi 2025]. It is also challenging to interpret UMAP distances for cluster analysis due to the non-linear transformation [Diaz-Papkovich 2021].

We therefore adopted topic modeling (TM) as a dimensionality-reduction and feature-construction step prior to unsupervised clustering of patients. TM is an unsupervised ML approach originally developed for document analysis, which is well suited for capturing co-occurrence structure (collinearity) from binary or count data by measuring and optimizing topic coherence (intra-topic similarity) and data likelihood (inter-topic distance) among the topics. When applied to a patient-symptom binary matrix, TM yields a concise set of interpretable symptom “topics” for downstream cluster analysis.

The proposed approach was assessed by correlating the clusters, endotypes, and severity scores identified by the machine learning analysis with known EQ-5D/EQ-VAS (EuroQol Visual Analogue Scale [Feng 2013]) scores, pre-existing conditions, and available antibody assay data. EQ-5D [Delvin 2020] is a self-reported questionnaire that assesses health across 5 categories: mobility, self-care, usual activities, pain/discomfort, and anxiety/depression. The accompanying EQ VAS reflects how a patient feels about their health overall on a scale of 0 to 100 (worst possible to best possible), which provides a continuous variable that is not limited by the 1, 3, and 5 ranking of the standard EQ-5D questions. Patient clusters are defined by symptom signatures. Endotypes are defined by organ systems involved. We connected symptoms to organ systems using curated symptom-to-organ mappings [Parotto 2023] in order to tag cluster signatures and identify endotypes. Clusters were merged hierarchically by shared symptom significance, and therefore, organ-system involvement. Each endotype was described by its top three organ systems. Cluster-level severity was quantified as the sum of topic proportions across symptoms for cluster members, after which an elbow method is used to discretize severity into qualitative levels (mild, moderate, severe). This symptom-based severity is interpretable in terms of signature symptoms and organ systems and, therefore, complements generic health-status scores (e.g., EQ-5D/EQ-VAS) for mechanistic and biomarker studies.

As clinical cohorts differ in enrollment criteria and COVID-specific case mix, we analyzed each cohort separately before integrating the results across cohorts. Our primary datasets included a walk-in clinic cohort (ISMMS), a longitudinal cohort spanning 4 years (UCSF LIINC), and a case-control study (Cardiff). We also included a cohort of 60 long COVID patients (Emory) with immunological assay data to validate severity predictions. Because the ISMMS cohort does not include inclusion or exclusion criteria, we merged Emory cohort into ISMMS (labeled as ISMMS+Emory) to assess robustness and cross-cohort reproducibility of patient groups, an out-of-cohort validation rarely performed in prior studies.

Figure 1 summarizes the data analytical workflow (Figure 1A) and illustrates its input (Figure 1B: a binary patient by symptom matrix) and outputs (Figure 1C-F: clusters, organ systems, endotypes, and severity groups). For each patient, our pipeline yielded four outputs: cluster assignment, endotype, continuous severity score, and discrete severity level. These outputs enabled cluster-specific biomarker association, recovery/worsening prediction, and cross-cohort comparison. All analysis code, data processing scripts, and figure generation routines are available in the project GitHub repository.

**Figure 1.**
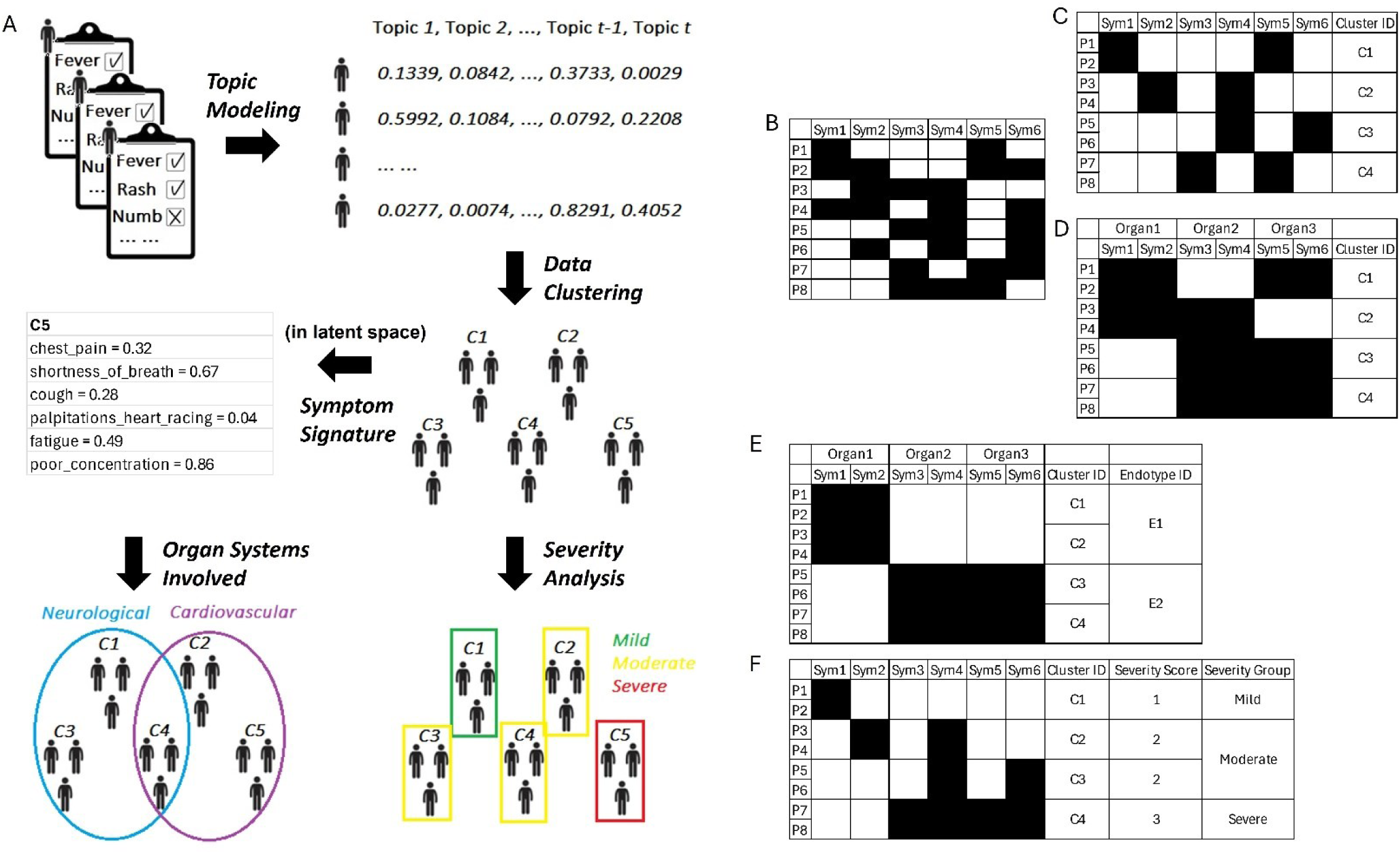
A schematic representation of the topic modeling-based symptoms analysis pipeline. **(A)** The input patient-by-symptom binary (presence or absence of a symptom) table is transformed into a patient-by-topic table by topic modeling so that unsupervised clustering analysis can be applied to identify patient clusters. For each cluster, we output the top six symptoms with the largest topic scores as the signature of the patient cluster. The clusters are grouped into multiple endotypes by mapping symptom signatures to related organ systems, followed by hierarchical agglomerative clustering. Each cluster is distinct but has one or more tags of organ systems. In parallel, we calculate severity of each cluster by adding the topic proportion values of all patients across all symptoms in the cluster. Illustration of an input table of patient by symptom: symptom presence in black and absence in white **(B)**. Illustration of clustering results **(C)**, organ systems **(D)**, endotypes **(E)**, and severity groups **(F)** identified from the input table and the relationship among these concepts.

## 2. Study Cohorts

### 2.1 UCSF

The *Long-term Impact of Infection with Novel Coronavirus (LIINC)* study is an ongoing University of California, San Francisco (UCSF)-based research program that evaluates the long-term health effects of COVID-19 [Peluso 2021]. Participants are recruited through self-referral or via a clinician. Any adult with a confirmed positive SARS-CoV-2 test is eligible. The study is not limited to those who believe themselves to have long COVID. The primary time variable was defined as the number of days between the first positive test and the date of survey completion.

Participants completed an initial survey during their first visit (baseline) and were invited to return approximately every 3 months for follow-up assessments. All questionnaires were administered by professional research staff in the context of a structured clinical interview. Only symptoms that were new or worse since the onset of COVID-19 and were not clearly attributable to another medical condition were attributed to long COVID. At the time of the data lock, LIINC had enrolled over 700 individuals and collected more than 2,000 surveys over 4 years, with each participant completing up to 12 surveys. After applying quality-control and completeness filters, 669 participants were included in our analysis. Surveys captured binary responses for 32 long COVID-attributed symptoms, the prevalence of pre-existing comorbidities (e.g., diabetes), demographics (e.g., sex, age), hospitalization status during acute COVID-19 (e.g., oxygen use, ICU admission, ventilator support), and quality of life using EQ-5D scores.

### 2.2 ISMMS

At the time of data analysis, the ISMMS clinic contributed data collected from over 1,200 patients, each of whom completed an initial survey and up to two follow-ups, totaling more than 5,000 surveys. Surveys with missing symptom reporting data were excluded, and full responses from initial visits were prioritized. This filtering yielded 615 participants for analysis. All participants met NASEM criteria for long COVID. In addition to symptom data derived from surveys, a committee from the ISMMS clinic attributed each of the symptoms measured to nine symptom subtypes associated with different organ systems or functions: neurological, sensory, cognitive, hormonal, musculoskeletal, digestive, temperature regulation, pulmonary, and cardiovascular [Parotto 2023].

### 2.3 Emory

The Emory long-COVID clinic enrolled 61 patients between January 2021 and February 2024, collecting binary symptom data, information on pre-existing conditions, and immunological measurements. These included MENSA (Media Enriched with Newly Synthesized Antibodies; [Haddad 2024]) and serum antibody assays. Sixty patients with complete clinical data were included in the present analysis.

### 2.4 Cardiff

The Cardiff case-control study collected symptom data along with demographic information from 317 participants at the time of analysis, consisting of 215 long COVID cases and 102 controls, all of whom had a clearly defined episode of acute COVID-19 accompanied by positive molecular test for SARS-CoV-2. Participants were enrolled between February 2022 and October 2023 in the United Kingdom. Other cohort details have been described previously [Baillie 2024, Gao 2025].

### 2.5 Data summary

Unique features of each cohort, the number of participants included, and the self-reported sex distributions are summarized in **Table 1**.

**Table 1.**
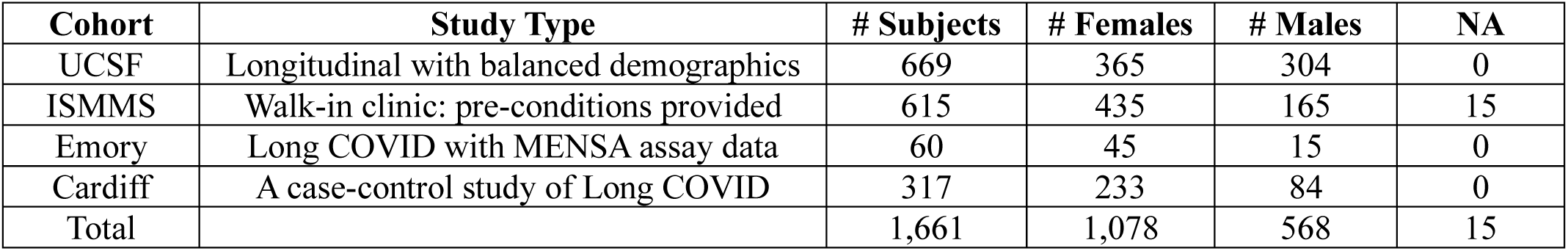
Cohort demographics and participant enrollment strategy.

| **Cohort** | **Study Type** | **# Subjects** | **# Females** | **# Males** | **NA** |
| --- | --- | --- | --- | --- | --- |
| UCSF | Longitudinal with balanced demographics | 669 | 365 | 304 | 0 |
| ISMMS | Walk-in clinic: pre-conditions provided | 615 | 435 | 165 | 15 |
| Emory | Long COVID with MENSA assay data | 60 | 45 | 15 | 0 |
| Cardiff | A case-control study of Long COVID | 317 | 233 | 84 | 0 |
| Total |  | 1,661 | 1,078 | 568 | 15 |

## 3. Results

Previous research already shows that a linear transformation of symptom features such as PCA does not help identify symptom clusters [Fresi 2025]. Compared with nonlinear embedding approaches such as UMAP which can be difficult to explain, topics identified in topic modeling (TM) are a weighted representation of original features. The topics can be mapped back to the original symptoms for interpretation. In pilot experiments, we demonstrated that TM produced finer and more precise symptom groupings than PCA or UMAP (**Supplementary Figure 1**). To address TM’s known challenges (non-determinism and topic-number selection), we generated multiple models to smooth the curves of topic coherence and data likelihood, followed by an ensemble criterion that balances the trade-off between these two metrics to choose the optimal topic count (**Supplementary Figure 2**).

To illustrate the range of outputs generated by our approach, results are organized by cohort. Section 3.1 focuses on symptom clusters identified in the ISMMS walk-in clinic. Section 3.2 reports severity scores derived from the combined ISMMS and Emory cohorts. Section 3.3 describes long COVID endotypes identified in the Cardiff case-control study. Section 3.4 examines longitudinal changes in symptoms and severity in the UCSF LIINC cohort. Section 3.5 presents a meta-analysis that integrates cohort-specific symptom clusters to identify endotypes and severity patterns across studies.

There are three ways this study groups patients from the topic modeling results: clusters, endotypes, and severity group. Clusters are derived from topic proportions (topic-by-patient), meaning patients with similar topic compositions are grouped together. Each topic represents a weighted set of symptom signatures defined in the topic matrix (topic-by-symptom). Conceptually, topics can be compared to genres used to describe books; books may belong to multiple genres (topics), and each genre is characterized by a particular, though not unique, set of words (symptoms). Patients are grouped into clusters, with the optimal number of clusters determined by the highest silhouette score, which measures how similar patients are within clusters and how distinct they are from other clusters. While clusters can be characterized by their most prevalent symptoms, we are also interested in understanding their broader organ-level associations. This is where endotypes come in. Endotypes group clusters based on the ISMMS-defined mapping of symptoms to symptom subtypes associated with different organ systems or functions [Parotto 2023], effectively summarizing clusters by their top organ associations and enabling cross-cohort comparisons. A limitation of this approach is that endotypes do not capture differences in symptom severity. To address this, we additionally grouped clusters by severity, which we defined using the cluster-by-symptom matrix. The cluster-by-symptom matrix is computed as the dot product of the topic-by-symptom and topic-by-cluster matrices, preserving the relative strength of association between clusters and specific symptoms.

### 3.1 Distinct symptom clusters identified in the ISMMS walk-in clinic cohort

Unlike cohorts recruited for clinical studies, ISMMS’s walk-in clinic did not apply inclusion or exclusion criteria, providing a broad overview of the heterogeneity and complexity of post-acute COVID symptoms. K-means clustering with K = 16 clusters achieved the highest silhouette score among all clustering methods and parameter choices (Figure 2A). For visualization, the topic proportions matrix was embedded with UMAP. Cluster quality was assessed using three criteria: (1) consistency with the UMAP layout (Figure 2A); (2) alignment of patient-level profiles with cluster-specific symptom signatures (Figure 2B); and (3) the presence of distinct symptom sets (i.e., signature symptoms) within each cluster (Figure 2C). All clusters satisfied these criteria. Cluster signatures were defined as the top six symptoms ranked by row-wise normalized cluster-by-symptom (i.e., normalized across clusters), representing the symptoms most specific to each cluster.

**Figure 2.**
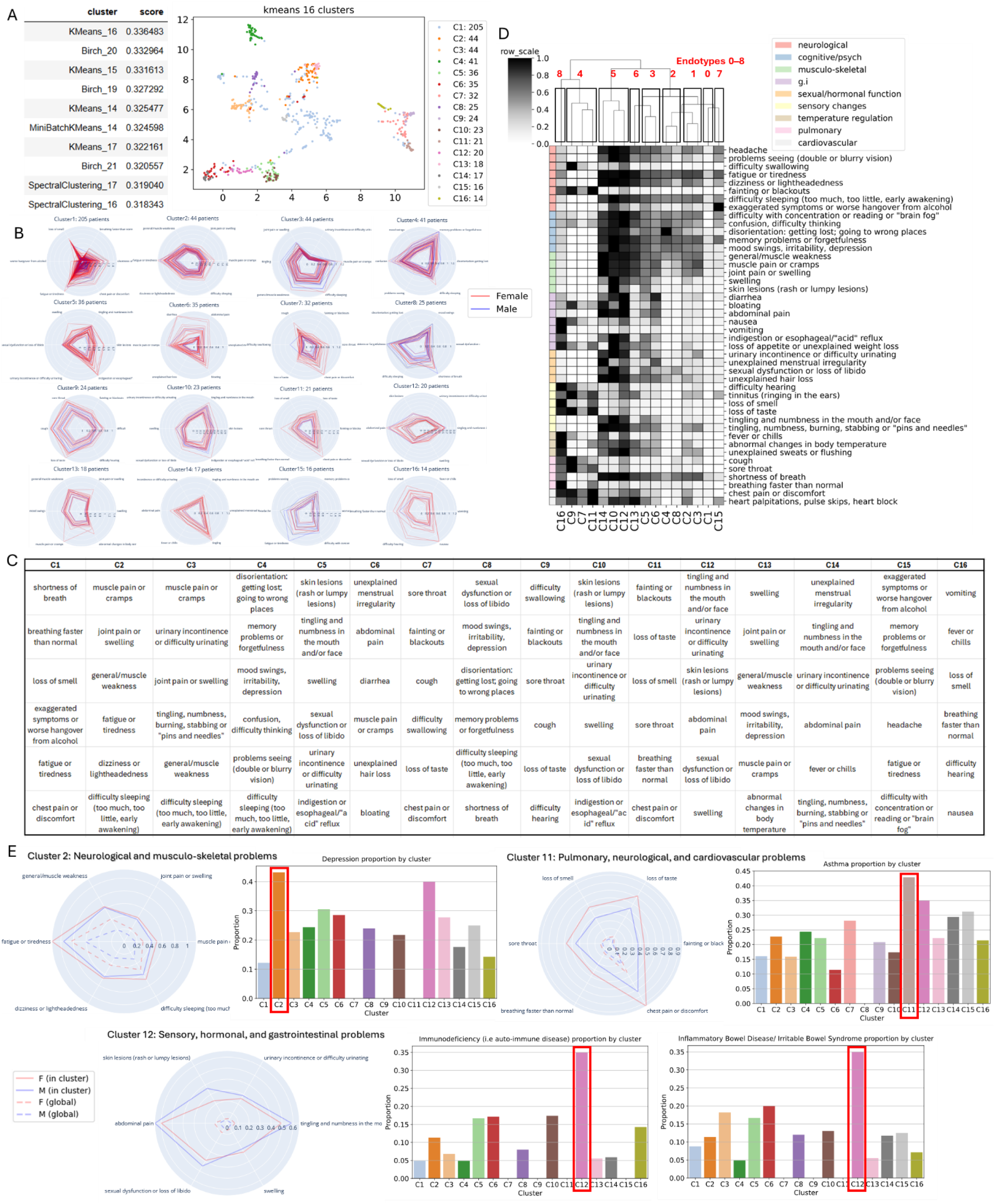
Identification of patient clusters, symptom signatures, and their association with pre-existing conditions from walk-in clinic cohort (ISMMS). (**A)** UMAP visualization of the 16 patient clusters. The table lists silhouette scores produced by different clustering algorithms and numbers of clusters. Each dot on the UMAP corresponds to one patient (n = 615) color-coded using cluster identifier (C1–C16). **(B)** Star charts visualizing top six signature symptoms identified in each cluster (C2 to C15; C1 is mild with few symptoms). Topic proportions in the patient-by-symptom table for each cluster were normalized before being visualized. Each ring corresponds to one patient, and all patients in each cluster are plotted together to demonstrate their consistent symptom profiles. Females are colored in red, and males are colored in blue. **(C)** Table listing symptom signatures of individual clusters. Each column corresponds to one cluster, and the top six symptoms are ranked based on their topic proportions, from high to low (Row 0 to Row 5). **(D)** Grayscale heatmap visualizing 16 symptom clusters merged into nine endotypes associated with predefined organ systems, following the dendrogram. Each row in the heatmap is a symptom and each column is a patient cluster. Rows/symptoms of the same organ system are approximated, with colors in the leftmost row indicating the organ system. A darker gray scale indicates a higher level of association between the cluster and the symptom, quantified using topic proportions. Cluster 1 (mild) corresponds to Endotype 0 (E0). Clusters with similar symptom signatures were grouped together using hierarchical clustering analysis. **(E)** Interpretation of each cluster. Each cluster is annotated with one or multiple involved organ systems based on its top six signature symptoms. Three examples (Clusters 2, 11, and 12) are shown. Four rings in each star chart are plotted to show the average topic proportion of all patients in a cluster for all females in the cluster (red line), all males in the cluster (blue line), all females in the cohort (red dotted line), and all males in the cohort (blue dotted line). For each pre-existing condition that is related to the organ systems, a bar chart was created to visualize the proportions of patients with the pre-existing condition across all patients in the cluster (one bar per cluster and the cluster of interest are highlighted).

To further organize symptom patterns, clusters were grouped into endotypes using hierarchical agglomerative clustering (Figure 2D). Endotypes were defined by organ system such that clusters with different symptom signatures but affecting the same organ systems were merged into a single endotype (See **Methods**). The dendrogram and top organ systems revealed nine endotypes (E0–E8), with severity represented by a grayscale heatmap. While E0 is clearly mild (white on heatmap), E5 displayed substantially greater severity than other endotypes (dark gray on heatmap) (Figure 2D).

Several clusters were interpretable in relation to pre-existing conditions (Figure 2E). For example, Cluster 2, characterized by joint pain or swelling, muscle pain or cramps, difficulty sleeping, dizziness or lightheadedness, fatigue, and generalized weakness, included 43% of patients with self-reported depression. Cluster 11, a pulmonary endotype, contained a sizeable proportion of patients with asthma (43%). Cluster 12, the most severe cluster based on the heatmap (Figure 2D), was marked by sensory, hormonal, and gastrointestinal problems; 35% of patients in this cluster reported autoimmune diseases or irritable bowel syndrome.

### 3.2 Correlation between ML-predicted symptom severity and SARS-CoV-2 antibodies in plasmablasts

Symptom severity was quantified as a cluster-specific feature by summing topic scores and proportions across all symptoms (Figure 3A). To validate the ML-predicted severity scores, we ran our endotyping pipeline on the ISMMS cohort twice, one alone and the other combined the ISMMS cohort (n = 615) with the Emory cohort (n = 60), which included serum antibody and MENSA (Media Enriched with Newly Synthesized Antibodies) assay data. The MENSA measures antibodies secreted exclusively by circulating plasmablasts, thereby capturing immune responses to recent infection or vaccination, in contrast to serum antibodies, which accumulate from past exposures [Haddad 2024]. This MENSA assay provides a snapshot of a patient’s immune status regarding exposures to viral proteins from SARS-CoV-2 and herpesviruses (Epstein Barr Virus: EBV), cytomegalovirus (CMV), and herpesvirus (HSV).

**Figure 3.**
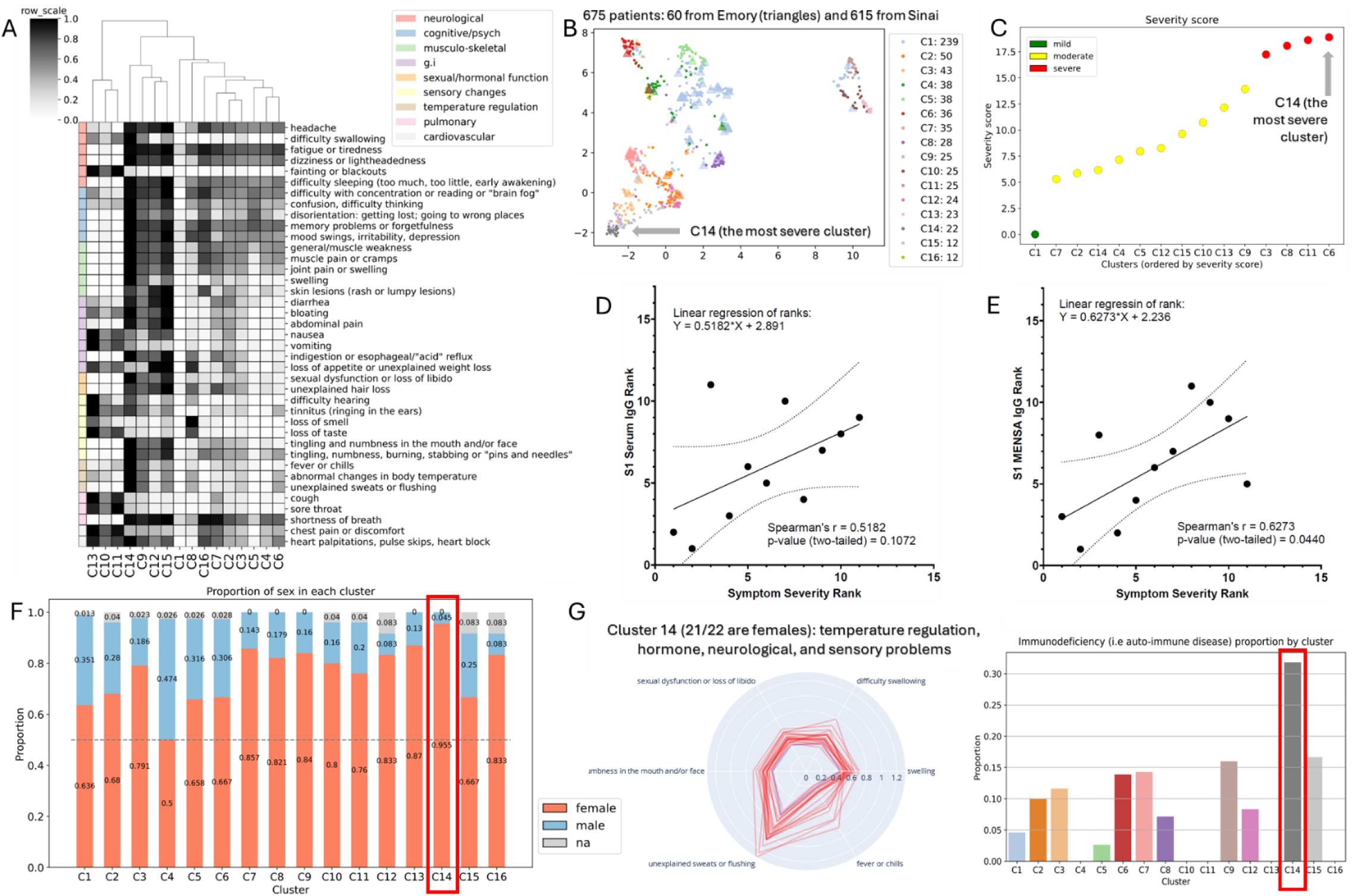
Interpretation of the severity of SARS-COV-2 infection and long COVID by association of symptom severity with antibody assay data (ISMMS+Emory cohorts). Surveys from ISMMS (n = 615) and Emory (n = 60) are merged as inputs into the topic modeling method. **(A)** Grayscale heatmap visualizing the cluster-by-symptom matrix. Each row is a symptom, and each column is a patient cluster. Symptoms are mapped to nine predefined organ systems. Three severity groups (mild: green; moderate: yellow; and severe: red) are identified and colored in the dendrogram based on a cluster-specific severity score, defined by sum of topic portions across all symptoms. **(B)** UMAP visualization of 16 patient clusters identified by the topic modeling-based method, ranked according to size. Cluster 14 (dark grey) is highlighted as the most severe patient group, which corresponds to Cluster 12 in the ISMMS-only analysis (Figure 2). **(C)** Identification of severity groups from a ranked view of cluster-severity plot. The 16 clusters (X-axis) were ranked based on the severity score (Y-axis). An elbow method was used to separate patient clusters into three groups: mild, moderate, and severe (also visualized in the dendrogram in Figure 3A). **(D, E)** Non-parametric rank-based analysis (X-axis: disease severity of patient clusters, ranked according to the sum of topic proportions across symptoms associated with the cluster; Y-axis: mean of antibody titers across all Emory samples within the cluster) showing correlations between topic modeling-predicted severity score and SARS-COV-2 S1 IgG in antibody binding assays (MENSA [Haddad 2024]). Spearman’s r = 0.5182 with a two-tailed p-value = 0.1072 and Spearman’s r = 0.6273 with a two-tailed p-value = 0.0440 were identified for Serum S1 and MENSA S1, respectively. The straight line was plotted from linear regression analyses with two curved lines showing the 95% confidence intervals. **(F)** Bar graph showing the proportions (Y-axis) of female (ochre) and male (blue) across clusters (X-axis). Cluster 14 is mostly female. **(G)** Star charts showing the 22 patients in Cluster 14, reporting symptoms associated with at least four types of organ systems. The bar chart highlights that 32% of the patients in this cluster have pre-existing condition with their immune systems.

Among the 16 symptom clusters (Figure 3B), three distinct severity groups, namely mild, moderate, and severe, were visually distinguishable based on gaps in the ML-predicted severity scores (Figure 3C). Linear regression analysis (Figure 3D-E) indicated likely positive correlations between severity scores and antibody responses against the SARS-CoV-2 S1 region of the spike glycoprotein (also the receptor binding domain and S2 regions; data not included). Nonparametric rank-based analyses confirmed these associations: the Spearman correlation between summed topic scores across symptoms reported in a cluster (the Emory samples were spread between 11 out of 16 clusters identified by the joint ISMMS+Emory analysis) and mean cluster-level IgG from MENSA was significant (Figure 3E, r = 0.6273, p = 0.0440). In contrast, pre-existing serum antibody titers showed a weaker association (Figure 3D, r = 0.5182, p = 0.1072).

Cluster 14 emerged as the most severe patient group, corresponding to Cluster 12 in Figure 2E. This cluster was predominantly female (21 of 22 patients F; Figure 3F) and characterized by symptoms related to temperature regulation, hormones, neurological disturbances, and sensory functions, consistent with pre-existing autoimmune conditions (Figure 3G). Adding 60 long COVID patients from the Emory cohort to the ISMMS dataset did not alter the identification of this female-dominated severe cluster, showcasing the robustness of the approach.

### 3.3 Identification of symptom clusters, organ-system endotypes, and severity groups in Long COVID

The Cardiff study employed a case–control design including 215 long COVID cases and 102 healthy convalescent controls (n = 317). Clustering analysis identified 16 symptom clusters. In a comparison with the ISMMS results, Cluster 1 in Cardiff corresponded primarily to controls, whereas Cluster 1 in ISMMS represented patients with fewer symptoms.

Based on signature symptoms, nine endotypes (E0–E8) were identified in the Cardiff cohort (Figure 4A). Organ systems associated with each endotype are summarized in Figure 4B. Six major systems were involved: gastrointestinal (E3, E5), pulmonary (E4, E7), cardiovascular (E1), musculoskeletal (E2, E3), sensory (E2, E5), and neurological (E2, E4). Among these, E3 and E6 represented the most severe endotypes: E6 was characterized by pulmonary and temperature regulation problems, while E3 combined gastrointestinal and musculoskeletal symptoms.

**Figure 4.**
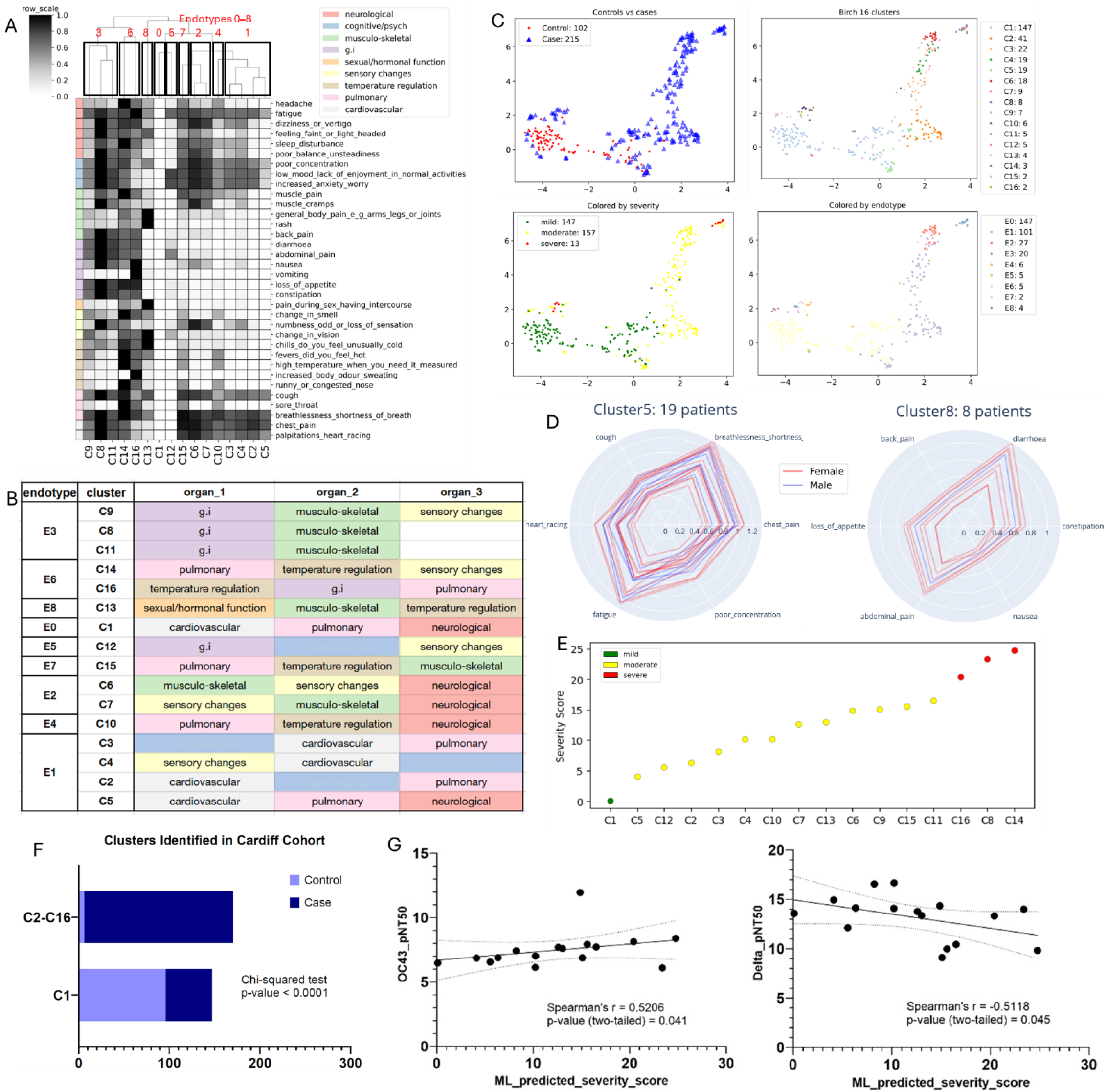
Identification of long COVID endotypes in a case-control study (Cardiff). **(A)** Grayscale heatmap showing 16 patient clusters (columns; C1 to C16) and nine endotypes (dendrogram; E0 to E8) identified in the Cardiff cohort (n=317, including 215 long COVID cases and 102 healthy convalescent controls) across 34 symptoms (rows; organized into nine pre-defined organ systems). **(B)** Table listing cluster members of endotypes and their signature organ systems (up to three included and colored as in Figure 4A). **(C)** UMAP visualization of patients, colored based on disease phenotype (upper left), symptom clusters (upper right), endotype (lower left), and severity (lower right). **(D)** Star charts showing two symptom clusters of interest. Cluster 5 incorporates cardiovascular and pulmonary symptoms, including chest pain, shortness of breath, and heart “racing”. Cluster 8 is a severe cluster with gastrointestinal symptoms, including diarrhea, nausea, and loss of appetite. Individual patients in the same cluster have consistent profiles across the signature symptoms. **(E)** Identification of severity groups from a ranked view of cluster-severity plot. The 16 clusters (X-axis) were ranked based on the severity score (Y-axis). An elbow method was used to separate the patient clusters into three groups: mild (green), moderate (yellow), and severe (red). **(F)** Bar graph showing significant overlap between clustering results in cases and controls (p-value < 0.0001, Chi-squared test). Cluster 1 (mild with few symptoms reported) includes controls and cases with mild symptoms. Clusters 2–16 correspond to cases with moderate and severe symptoms. **(G)** Linear regression analyses visualizing correlations between virus neutralization and severity score (X-axis: rank of patient clusters based on the sum of topic proportions across all symptoms. Y-axis: mean of antibody titer values across all patients in the same cluster). Neutralization of OC43, a betacoronavirus, correlates directly with long COVID severity (Spearman’s r = 0.5206, p-value = 0.041), whereas neutralization of the Delta variant correlates inversely with long COVID severity (Spearman’s r = −0.5118, p-value = 0.045).

Symptom clusters, endotypes, and severity groups were overlaid with case–control status for visual comparison (Figure 4C). The UMAP based on topic scores revealed two major “islands” corresponding to cases and controls. Clusters, endotypes, and severity groups further partitioned these islands, providing finer stratification consistent with the UMAP layout. Notably, some cases exhibited mild or few symptoms resembling convalescent controls, while a subset of controls reported symptoms similar to those observed in cases (Figure 4C, upper left). E3 and E6, although both severe, were positioned at opposite ends of the UMAP space (Figure 4C, bottom left).

Representative clusters illustrate the diversity of endotypes. Cluster 5 (E1: pulmonary/cardiovascular endotype) included 19 patients with consistent profiles of chest pain, shortness of breath, and heart racing (Figure 4D, left). Cluster 8 (E3: gastrointestinal endotype) comprised patients with diarrhea, abdominal pain, loss of appetite, constipation, and nausea (Figure 4D, right). Within the gastrointestinal endotype (E3), clusters 8, 9, and 11 all presented gastrointestinal symptoms but differed in severity levels (Figure 4E).

Comparison of Cluster 1 with controls using a contingency table (C1 vs. non-C1 against case vs. control status) demonstrated strong concordance between ML-derived clustering and the clinical diagnosis of long COVID (Chi-squared test, p < 0.0001; Figure 4F). Neutralization assay data further supported these findings. Specifically, these ML-predicted severity scores correlated modestly but significantly with antibody responses: they were positively associated with blood titers against OC43, a common coronavirus (Spearman’s r = 0.52, p = 0.041), and negatively associated with titers against SARS-CoV-2 Delta (Spearman’s r = −0.51, p = 0.045).

### 3.4 Identification of symptom changes and recovered/worsened subgroups in a longitudinal study

The UCSF LIINC cohort collected symptom data following SARS-CoV-2 infection over a 4-year longitudinal period (Figure 6C). The cohort included both fully recovered and long COVID patients; a subset had enrolled during the acute phase of COVID-19 and were followed longitudinally. Within LIINC, long COVID is defined as the presence of at least one COVID-attributed symptom that is present in the post-acute phase of illness, consistent with the NASEM case definition [NASEM 2024]. Therefore, we used Day 85 as the threshold to separate acute and post-acute phases from long COVID (Figure 6C).

Using symptoms reported in the first survey of each patient (n = 669), our topic modeling approach identified 15 symptom clusters (Figure 5A). UMAP visualization was consistent with the clustering results, revealing one large cluster of patients with few symptoms (Cluster 1, n = 251) alongside 14 distinct symptomatic clusters. Surveys from the first visit of each patient were included in the analysis, which are divided into three stages and visualized separately: acute (Day 0-30), early post-acute (Day 31-90), and long COVID (after Day 90). Arrows were used to highlight the corresponding symptom clusters and symptom severity in each phase: acute (Cluster 2: Moderate; Temperature regulation, Pulmonary), early post-acute (Cluster 1: Mild; Musculo-skeletal, Pulmonary), and long COVID (Clusters 3, 6, 8, 11: Severe, Neurological, Hormonal function, Sensory changes, and Musculo-skeletal). Connecting these symptom clusters across the three phases/UMAPs discloses a trajectory of symptom changes from acute to long COVID. Heterogeneity of symptoms and organ systems associated with long COVID is clearly higher than the earlier phases, while the symptom severity at infection becomes milder after the acute phase before it gets worse at long COVID. From the 15 symptom clusters, 12 endotypes were derived (Figure 5B), suggesting greater heterogeneity of affected organ systems in the LIINC cohort compared to the Cardiff or ISMMS cohorts. As in the latter two cohorts, three severity levels, namely mild, moderate, and severe, were also evident in the LIINC cohort (Figure 5C).

**Figure 5.**
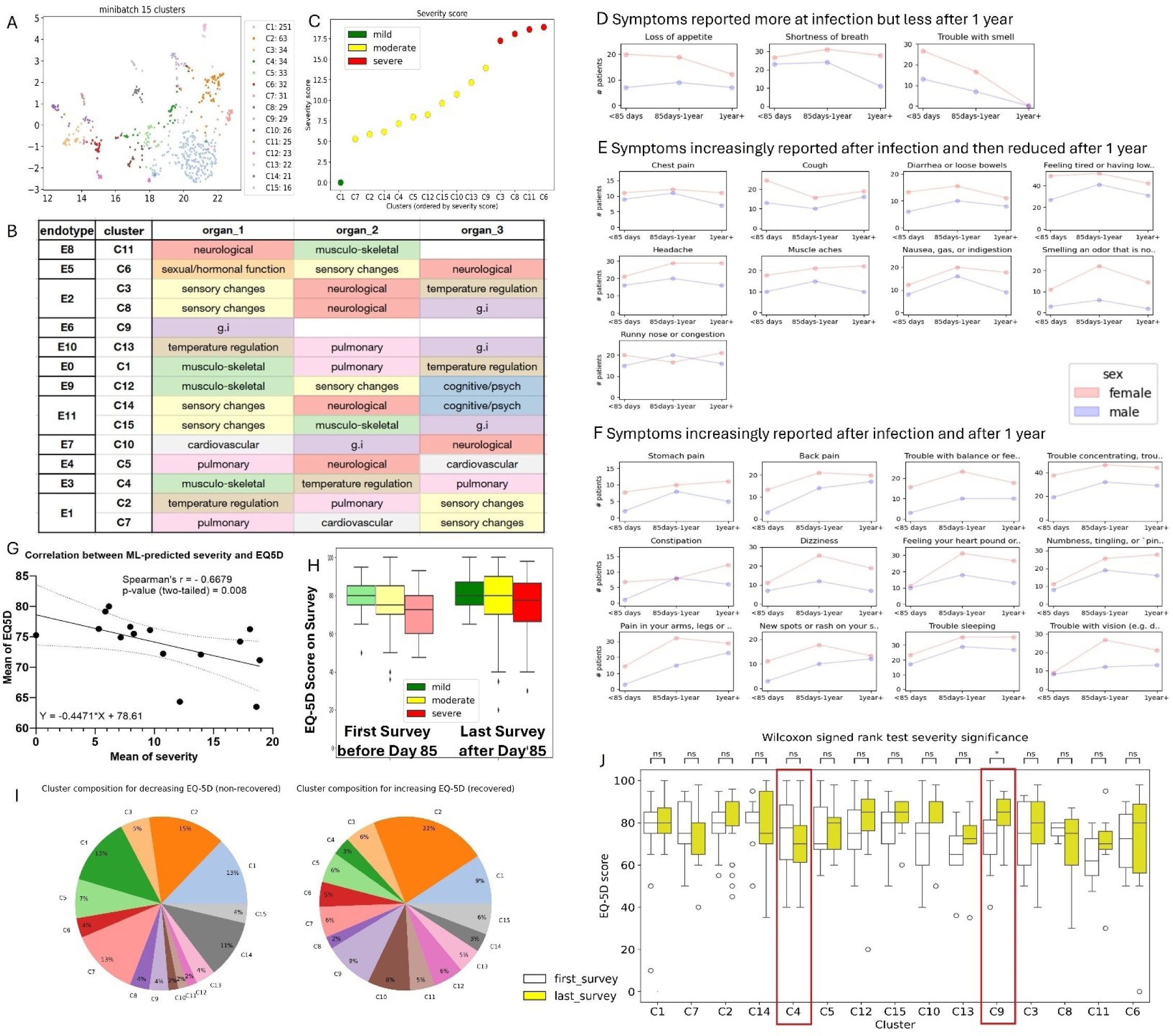
Identification of symptom clusters and severity changes in a longitudinal study (LIINC cohort). **(A)** UMAP visualization of 15 symptom clusters (C1 to C15) identified from first-visit surveys after SARS-COV-2 infection (n=669 in total). **(B)** Table of endotypes (E0 to E11) and their signature organ systems (up to three included). **(C)** Identification of severity groups from a ranked view of cluster-severity plot. The 15 clusters (X-axis) were ranked based on the severity score (Y-axis). An elbow method was used to separate the patient clusters into three groups: mild (green), moderate (yellow), and severe (red). **(D–F)** Line graphs showing the frequency of self-reported symptoms at three time points after SARS-COV-2 infection: 0 to 85 days, 85 days to 1 year, and after1 year. Some symptoms disappeared after 1 year (**D**), while others did not alleviate (**E**) or even became worse (**F**). **(G)** Linear regression analysis visualizing the correlation between EQ-5D and severity score (X-axis: rank of patient clusters based on sum of topic proportions across all symptoms; Y-axis: mean of EQ-5D across all patients in the same cluster). ML-predicted severity correlated inversely with EQ-5D (Spearman’s r = −0.6679, p-value = 0.008). **(H)** Box plot showing the change in health status (quantified by EQ-5D) for patients (n = 162) who completed surveys at both before and after day 85 post SARS-COV-2 infection. Severity groups were originally identified based on first surveys (n=669): mild in green, moderate in yellow, and severe in red. The first available survey before Day 85 and the last survey after Day 85 were used, when multiple surveys were available. **(I)** Pie charts showing the distribution of symptom clusters (C1–C15) among non-recovered patients (left; n = 55; EQ-5D decreased after day 85) and recovering patients (right; n = 87; EQ-5D increased after day 85), expressed as percentages. The percentage of Cluster 4 (Endotype 3, musculo-skeletal and other problems) is large for non-recovered but small for recovered patients, whereas the percentage of Cluster 9 (Endotype 6, gastrointestinal problems only) is large for recovered but small for non-recovered. **(J)** Box plot confirming significant changes in EQ-5D scores for patient clusters between the first survey before day 85 and the last survey after day 85 (X-axis: EQ-5D scores in the past week of the same visit; Y-axis: 15 clusters ranked by severity in the first survey). The patients in Cluster 4 reported lower EQ-5D scores after day 85, whereas those in Cluster 9 have significantly increased EQ-5D scores (p-value < 0.05, Wilcoxon signed rank test).

Among the 669 patients, 162 had surveys spanning ≥1 year. For this subgroup, we examined symptom frequency across three stages: before day 85, between day 85 and 1 year, and beyond 1 year. Based on frequency trajectories (with male and female patients analyzed separately), symptoms were classified into three groups: (1) Acute-resolving symptoms – present during acute infection but nearly absent after 1 year (e.g., cough, fever, loss of appetite, impaired smell; Figure 5D); (2) Persistent but attenuated symptoms – increased after infection, declined over time, but remained at low levels beyond 1 year (e.g., fatigue, stomach pain, chest pain, diarrhea, headache, shortness of breath, dizziness, cramps, muscle aches; Figure 5E); (3) Progressive symptoms – worsened compared to acute infection and remained elevated after 1 year (e.g., back pain, chills, balance problems, joint pain, palpitations, concentration difficulty, numbness, rash, vision problems, sleep disturbance; Figure 5F).

To validate ML-predicted severity, EQ-5D scores were plotted across severity groups before and after Day 85 (Figure 5H). Most patients demonstrated partial recovery after day 85, reflected by improved EQ-5D values. Severity levels are consistent with EQ-5D levels (Figure 5H). ML-predicted severity scores were negatively correlated with EQ-5D (Spearman’s r = −0.67, p = 0.008; Figure 5G), providing partial validation.

Based on the EQ-5D trajectories, patients were stratified into *recovered* (increasing EQ-5D) and *non-recovered* groups (decreasing EQ-5D). To assess which clusters were more likely to recover, we compared the cluster composition of these groups (Figure 5I). Cluster 4 (E3: moderate musculoskeletal and temperature regulation with pulmonary involvement) was enriched among non-recovered patients, while Cluster 9 (E6: gastrointestinal problems) and Cluster 10 (E7: cardiovascular problems) were more common in recovered patients. EQ-5D changes between the first surveys (before day 85) and last surveys (after day 85) confirmed these trends (Figure 5J). Of note, Cluster 9 showed significant improvement (p < 0.05), suggesting substantial recovery of gastrointestinal-only cases.

The clustering results shown in Figure 5A were based on the first surveys completed by 669 patients. Of these, 222 surveys were completed during the acute phase (Day 0–30), 235 during the early post-acute phase (Day 31–90), and 212 during the long COVID phase (Figure 6A). Visualizing the 15 clusters side by side across the three phases (Figure 6A) enables assessment of changes in symptom severity over time. Clusters C1 (mild) and C2 (moderate) were the most prevalent during the acute phase (Figure 6A**, left**). In the early post-acute phase, the size of C1 increased while the size of C2 decreased, indicating recovery in a subset of patients (Figure 6A**, middle**). In contrast, during the long COVID phase, both symptom severity and the heterogeneity of involved organ systems increased (Figure 6A**, right**). Neurological, hormonal, and sensory symptoms emerged predominantly during the long COVID phase. Notably, the sizes of all four severe clusters (C3, C6, C8, and C11) increased substantially after Day 90 (Figure 6B). Together, these observations suggest two divergent symptom trajectories: while some patients recover following acute infection, others progress toward severe long COVID symptoms.

**Figure 6.**
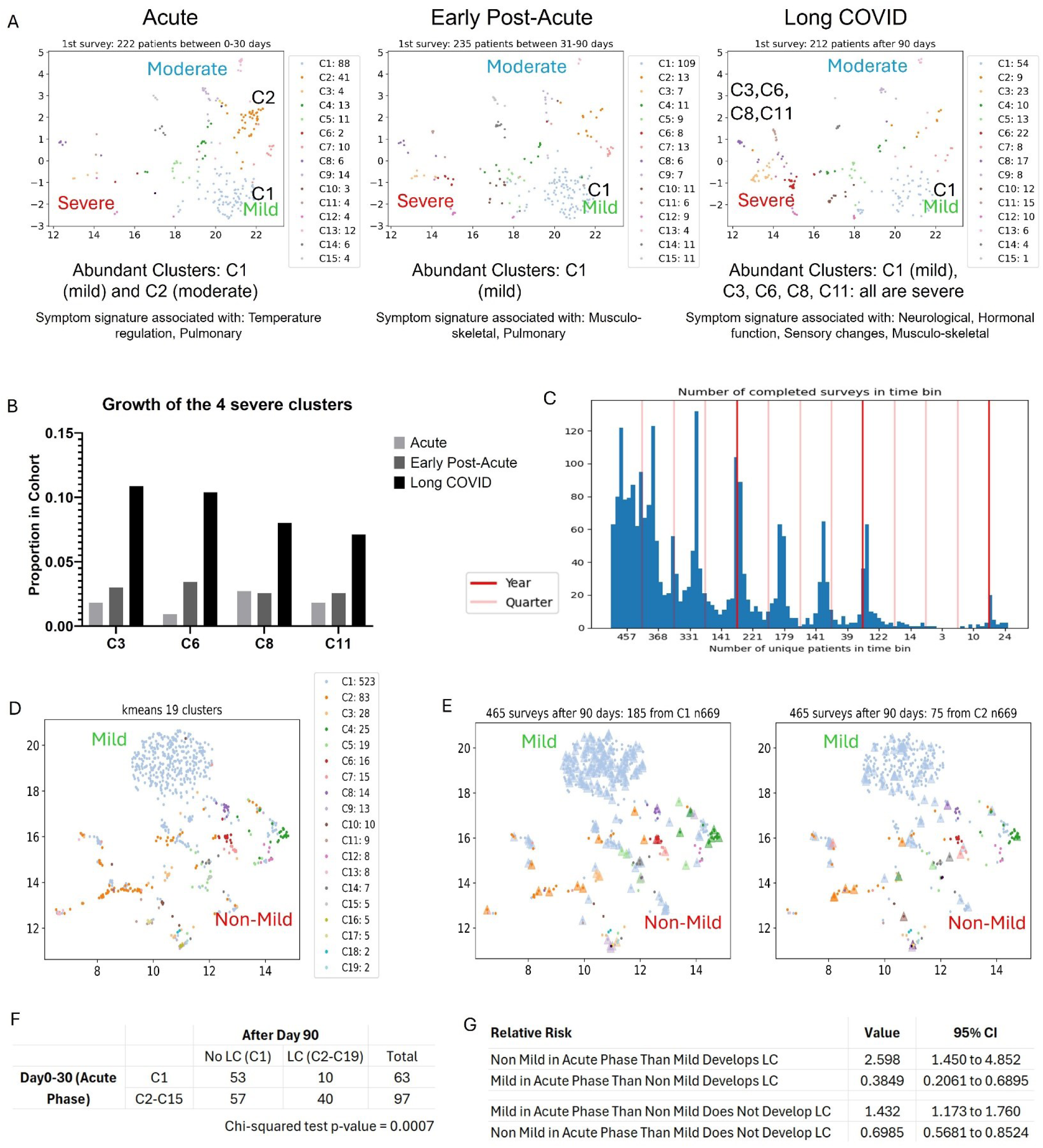
Identification of longitudinal trajectories of symptom clusters (LIINC cohort). **(A)** UMAP visualization of 15 symptom clusters (C1 to C15) identified from first-visit surveys after SARS-COV-2 infection (n=669 in total), organized based on “*Days into Infection*”: acute (Day 0 to 30, n = 222), early post-acute (Day 31 to 90, n=235), and long COVID (after Day 90, n = 212). C1 and C2 are the two most abundant clusters in the acute phase. Two trajectories of symptom clusters in the acute phase can be seen across these three UMAPs: evolving to long COVID (increasing size of the severe group in the long COVID phase) or recovering (increasing size of the mild group in early post-acute phase). Heterogeneity of organ systems involved and increasing severity are observed in the long COVID phase **(B)**. **(C)** Histogram showing the distribution of surveys across time (X-axis: 3-year duration, yearly separated by red lines). At each visit per quarter (pink line), there is a peak for the number of completed surveys. Surveys of 457 patients (222 between Day 0-30, and 235 between Day 31-90) were completed in the first quarter. **(D)** A longitudinal study: 19 clusters were identified from questionnaires across all time of the 222 patients who have completed surveys in both acute and long COVID phases. Distribution of surveys from patients in original clusters C1 **(E)** and C2 **(F)** in the long COVID phase. Majority of C1 surveys remain mild after Day 90, while many of C2 surveys evolve to long COVID after Day 90. **(F)** Among the 63 patients in the original C1 cluster who have completed surveys in both acute and long COVID phases, 53 remain mild and 10 develop moderate to severe symptoms. In contrast, 40 of the 97 patients that are not mild (C2-C15) in the acute phase still have moderate to severe symptoms after Day 90. Chi-squared test p-value (0.0007) shows there is significant association between the severity in the acute phase and the long COVID phase. **(G)** Individuals with symptoms in clusters C2 to C15 have 2.6 times (relative risk value = 2.598) the risk of developing long COVID compared with individuals with mild symptoms. Reciprocal relative risks and 95% confidence intervals are also calculated.

To further investigate longitudinal patterns, we focused on the 222 patients who completed surveys during both the acute and long COVID phases. Surveys from all available time points for these patients were included in a separate clustering analysis, which identified 19 clusters (Figure 6D). We then examined changes in symptom cluster membership after Day 90. Most patients in C1 remained mild after Day 90, whereas many patients in C2 progressed to moderate or severe symptom clusters (Figure 6E). To quantitatively assess changes of patient numbers between phases, we performed a chi-squared test, which showed a significant association between acute-phase severity and long COVID severity (*p = 0.0007*). Specifically, 40 of 97 patients with moderate or severe symptoms during the acute phase (C2–C15) developed long COVID, whereas only 10 of 63 patients with mild symptoms during the acute phase developed moderate or severe symptoms after Day 90 (Figure 6F). Risk factor analysis indicated that individuals with non-mild symptoms during the acute phase had a 2.598-fold higher risk (95% confidence interval: 1.450–4.852) of developing moderate or severe long COVID compared with individuals with mild acute symptoms. Conversely, individuals with mild symptoms during the acute phase had a 1.432-fold higher likelihood (95% confidence interval: 1.173–1.760) of remaining mild after Day 90 compared with those with non-mild acute symptoms (Figure 6G).

### 3.5 Meta-analysis defines global severity scores and identifies cross-cohort endotypes

To compare characteristics across cohorts, we merged cohort-specific clusters using a common set of 22 symptoms (mapping in Supplementary Table *data_dictionary*). Column-wise normalization of topic proportions (i.e., normalized across symptoms/organ systems) identified differences between cohorts in terms of organ system problems being reported (Figure 7A, numbers highlighted in red): (1) gastrointestinal problems are a distinct group in Cardiff and UCSF; (2) pulmonary problems are obvious in ISMMS, Emory and UCSF; (3) musculoskeletal problems are frequently reported in all three cohorts; (4) cardiovascular problems are clearly associated with Cardiff patients; (5) sexual/hormonal function problems are typical in ISMMS and Emory patients.

**Figure 7.**
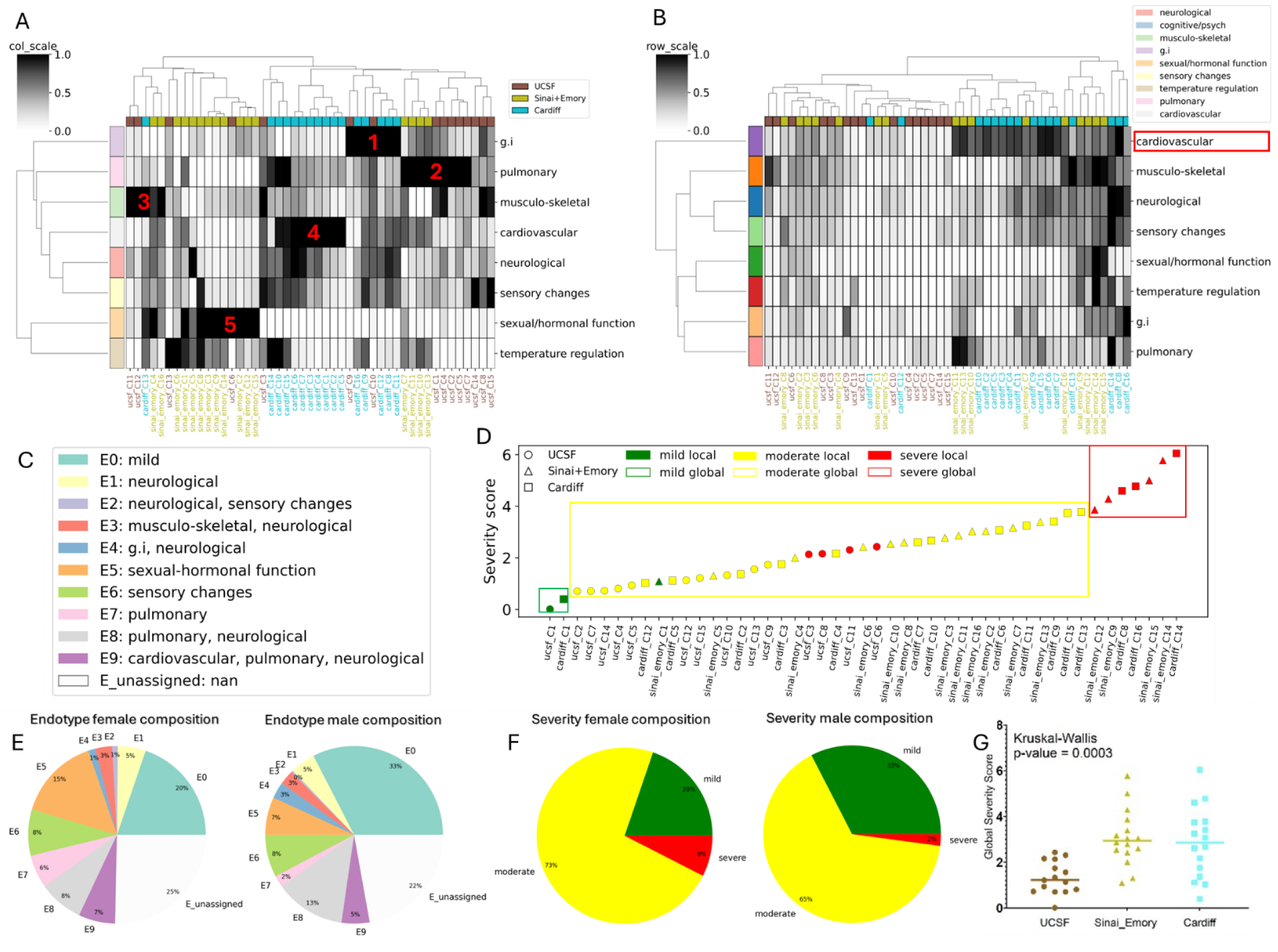
Comparison of endotype characteristics and symptom severity levels across cohorts. **(A)** Grayscale heatmap visualizing associations between patient clusters across sites (X-axis) and organ systems involved (Y-axis) using column-based normalization. Five distinct organ system-specific meta-clusters are present in different cohorts: (1) gastrointestinal problems reported by Cardiff and UCSF; (2) pulmonary problems reported by ISMMS+Emory and UCSF; (3) musculoskeletal problems by all sites; (4) cardiovascular problems by Cardiff; and (5) hormonal problems by ISMMS+Emory and UCSF. **(B)** Grayscale heatmap visualizing associations between patient clusters across sites (X-axis) and organ systems involved (Y-axis) using row-based normalization. The cardiovascular endotype reported by Cardiff is distant from the others in the organ-system dendrogram. Neurological and sensory problems are the closest organ systems. **(C)** Clusters identified in individual cohorts (47 clusters in total) merged into ten endotypes based on a common set of symptoms reported across cohorts. Clusters identified from cohort-specific symptoms, which cannot be assigned to any of the ten endotypes, are grouped into E_unassigned. **(D)** Identification of global severity levels along patient clusters in different cohorts by normalizing cluster-specific/local severity scores across cohorts on common sets of symptoms. The 47 clusters (X-axis) include two mild (green), 38 moderate (yellow), and seven severe (red). None of the UCSF clusters (circles) were globally severe compared with those in ISMMS+Emory (triangles) and Cardiff (squares). None of the ISMMS+Emory clusters (triangles) were globally mild. **(E)** Pie charts visualizing female (left) and male composition (right) for endotypes. Female contains a larger percentage of patients with sexual-hormonal problems (E5) than male. **(F)** Pie charts visualizing female (left) and male composition (right) for severity levels. Female contains a larger percentage of moderate and severe cases than male. **(G)** Symptom severity reported by UCSF patients (first surveys used) is significantly smaller than that of ISMMS+Emory or Cardiff (p-value = 0.0003, non-parametric ANOVA). Each dot represents a patient cluster identified across cohorts (X-axis) with its global severity score (Y-axis).

We then explored relationships among organ systems using row-wise normalization of topic proportions across all patient clusters (Figure 7B). The dendrogram shows that cardiovascular problems are distant from (i.e., not often reported together with) other organ systems, whereas neurological and sensory systems are closely linked (i.e., often reported together). Both are close to musculoskeletal problems. Sexual/hormonal function and temperature regulation are frequently reported together, and gastrointestinal and pulmonary issues also appear related.

Based on common symptoms shared across cohorts, patient clusters per site were further merged into ten global endotypes (E0–E9, Figure 7C). Some original clusters could not be assigned to any global endotype (labeled *E_unassigned* in Figure 7C), meaning clusters across cohorts could not be merged. Recalculation of severity scores using the updated topic proportions produced a global severity map across cohorts (Figure 7D). UCSF and Cardiff’s original mild groups (both Cluster 1) remained mild, while Cluster 1 from ISMMS+Emory shifted to moderate severity, consistent with the fact that walk-in clinic patients typically present with symptoms, whereas Cluster 1 from Cardiff comprised mostly convalescent controls. Of note, the most severe clusters from UCSF reported less symptoms than those in Cardiff and ISMMS, likely because the first surveys post-acute-infection in the UCSF LIINC cohort were used, which captured only early symptoms.

These cohort studies were not randomized, and gender distributions were imbalanced. Accordingly, we examined the composition of endotypes and severity groups by sex, rather than comparing the raw numbers of males and females side-by-side (Figures 7E**–F**). Overall, females reported more symptoms than males, consequently occupying a smaller proportion of the mild group (E0) and a larger proportion of the severe group. Pulmonary problems were common in both sexes (E7 and E8), but males were more likely to report concurrent pulmonary and neurological symptoms (E8).

Differences in ML-predicted severity between cohorts were statistically significant (non-parametric one-way ANOVA, p = 0.0003, Figure 7G) and likely reflect differences in how the different cohorts were assembled. UCSF patients appeared to have less severe symptoms than patients in the other cohorts, likely due to its lower proportion of female participants relative to the other cohorts (**Table 1**).

## 4. Methods

Steps in our pipeline are described in Section 4.1. Methods used in each step are described in greater detail in Section 4.2.

### 4.1 General data analysis steps

**Step 1: Pre-process surveys.** Each questionnaire was converted into a binary patient-by-symptom matrix indicating the presence or absence of each symptom. Quality-control and completeness filters were applied before downstream analysis.

**Step 2: Select optimal number of topics.** The number of topics was determined by an ensemble criterion that combines topic coherence (measuring intra-topic semantic consistency) and held-out likelihood (model fit). The chosen topic count balances interpretability and statistical fit.

**Step 3: Transform features using topic modeling.** Topic modeling (PFA: an unsupervised machine learning approach) was applied to transform the binary patient-by-symptom table into a patient-by-topic matrix and topic-by-symptom matrix. Each topic is a weighted combination of symptoms representing a non-linear summary of symptom co-occurrence.

**Step 4: Cluster analysis and signature extraction.** The patient-by-topic matrix was clustered and evaluated using a variety of clustering algorithms, number of clusters K, and silhouette scores. The clustering solution with the highest silhouette score was selected. For each cluster, the top six signature symptoms, ranked by row-wise normalized topic scores, were reported.

**Step 5: Visual assessment with UMAP.** A UMAP embedding of the normalized patient-by-topic matrix was computed independently for visualization. The UMAP layout was used to visually confirm cluster separation and to overlay metadata (e.g., case/control, sex, severity groups).

**Step 6: Map to organ systems and define endotypes.** Each cluster’s top signature symptoms were mapped to general organ systems using the ISMMS predefined symptom-to-organ mapping [Parotto 2023]. Hierarchical agglomerative clustering of cluster-level symptom profiles was applied to merge clusters that involve similar organ systems into endotypes. Each endotype is defined by its top three organ systems with associations visualized in a grayscale heatmap. Given that a cluster is a group of patients with the same symptom signatures, an endotype is grouped clusters with similar symptom signatures.

**Step 7: Compute severity scores and define severity groups.** Cluster-level severity is calculated as the sum of the cluster’s topic proportions across symptoms (visualized as darker shades in the heatmap). An elbow method on the distribution of severity scores was applied to define discrete severity groups (mild, moderate, severe).

**Step 8: Association analyses.** Associated demographics (sex, age), clinical phenotypes (case vs. control, hospitalization), pre-existing conditions, and immunological assays (e.g., MENSA, serum antibodies) with clusters, endotypes, and severity groups using regression and correlation analyses.

### 4.2 Detailed step-specific methods

#### Pre-processing the surveys

Patients completed questionnaires on paper or virtually via a REDCap survey and database. Results without protected health information (PHI) were then exported into a table along with derived variables such as age, body mass index (BMI), and number of days since the first positive test for SARS-CoV-2. Self-reported presence or absence of symptoms formed the binary patient-by-symptom matrix used in topic modeling. Short-form representations of each question were used as column names, with a data dictionary containing the full descriptions. Each row encapsulated a single patient survey. Duplicate or incomplete entries were either removed (when 2 or more symptoms were missing) or imputed (when only 1 symptom is missing). Imputation was performed using the mean value of other symptoms within the same organ system.

Long COVID was defined by persistent symptoms beyond 3 months. We applied an 85-day cutoff to ensure the inclusion of surveys that were completed only a few days earlier than Day 90 cutoff. For the longitudinal analysis of EQ-5D scores (UCSF only), we required patients to have completed at least two surveys: one survey before day 85 and another survey after day 85, both including EQ-5D scores. Using these criteria, the UCSF longitudinal analysis included 162 patients with two surveys each. For the cross-sectional analysis, the survey time was recorded as the order of completion rather than days since infection. The topic modeling was applied to the first available survey of the patient (UCSF, ISMMS+Emory, Cardiff).

#### Topic modeling

Our approach to defining COVID-related endotypes was inspired by methods described previously [Zhang 2023]. We applied PyPDM’s Poisson Factor Analysis (PFA [Zhou 2012]), a probabilistic topic modeling algorithm, to binary symptom data to define latent patterns (topics) that characterize each patients’ symptoms. A model was generated for each cohort (UCSF, ISMMS+Emory, Cardiff). A joint topic model was generated from ISMMS and Emory data (referred to as ISMMS+Emory). The model input consisted of binary symptom data from the first completed survey and an optimized number of topics. The model produces two key outputs: a topic matrix (topic-by-symptom), which defines each topic by the relevance or frequency of associated symptoms, and a topic proportion matrix (topic-by-patient), quantifying the extent to which each topic contributes to an individual patient’s symptom profile. Topic modeling operates under the assumption that each patient’s symptom set is a combination of underlying latent topics (groups of symptoms). The idea of topic modeling is comparable to how documents can be described by combinations of subject topics in natural language processing. Because our binary symptom data is sparse compared to text documents, we used Poisson Factor Analysis (PFA [Gan 2015]) which was derived from the more popular Latent Dirichlet Allocation (LDA). The following equation is used to describe the model in PFA for symptom s, topic t, and patient p:

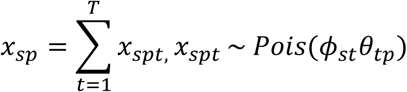

x_sp_ represents the observed count (patient p reporting symptom s). ϕ_st_ represents the factor loading matrix (topic matrix: topic-by-symptom). θ_tp_ represents the factor score matrix (topic proportions: topic-by-patient). The observed count (x_sp_) undergoes latent nonnegative matrix factorization (NMF) to decompose into the factor loading and score matrices.

The observed count is modeled with a Poisson distribution, which is used for modeling nonnegative integer count data such as the presence (1) or absence (0) of a symptom in a patient’s survey. ϕ_st_ is drawn from a Dirichlet prior, ensuring symptom weights are nonnegative and sum to one. θ_tp_ is drawn from a Gamma distribution, allowing for overdispersion and variability. To determine the optimal number of topics, we utilized a combined score based on PyDPM’s data likelihood [Zhou 2012] and Gensim’s topic coherence [Röder 2015] (https://radimrehurek.com/gensim/models/coherencemodel.html). Data likelihood measures how well the model fits the observed symptom data, while topic coherence evaluates the consistency of symptoms within each topic. Given the stochastic nature of topic modeling and its sensitivity to initialization, we ran the model ten times for each candidate topic number and averaged the results to stabilize performance metrics and reduce variance. Balancing topic coherence and data likelihood, this combined score ensures that the grouped symptoms are clinically interpretable. This approach enabled us to identify a robust number of topics for each model, which served as the foundation for downstream clustering and endotype definition, by maximizing the following score:

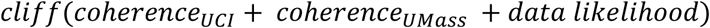

with data likelihood for P patients and S symptoms defined as:

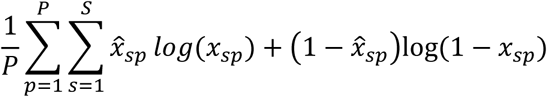

x_sp_ is defined above.

Topic coherences were defined as follows:

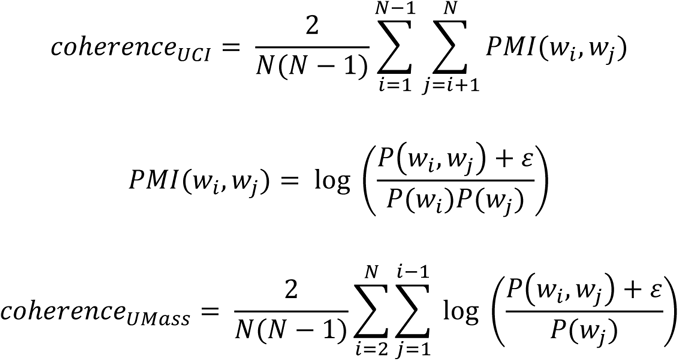

These topic coherence equations are calculated from a sliding window of word w and N top words. UCI coherence is based on the pointwise mutual information (PMI), where probabilities are based on word co-occurrence counts. UMass coherence is similar to PMI but the probabilities are based on reference documents. There are multiple ways to calculate coherence, so we incorporated both into our optimal number of topics calculation.

#### Clustering

Following topic modeling, each patient was represented by a topic proportion vector, capturing the degree to which each latent topic contributed to their overall profile. These topic-based representations were then used to cluster the patients. This step introduced a second key parameter: the number of patient clusters. To identify the optimal number of clusters and ensure that the resulting groups were distinct, we evaluated several scikit learn distance clustering algorithms and selected the final method based on overall performance measured via the scikit learn silhouette score, which quantifies how well each patient fits within its assigned cluster relative to all other clusters. The clustering methods assessed were *kmeans*, *minibatch kmeans*, *birch*, *spectral clustering*, *agglomerative clustering*, and *bisecting kmeans* (all methods available in module *sklearn.cluster*).

Average per sample silhouette scores evaluate cluster quality and balance intra-cluster and nearest-cluster distances. The clustering method and number of clusters that maximized the average silhouette score [Rousseeuw 1987] were chosen before downstream analysis. Clusters were labeled in descending order according to the number of patients (i.e. cluster 1 having the most patients). The silhouette score is defined as follows:

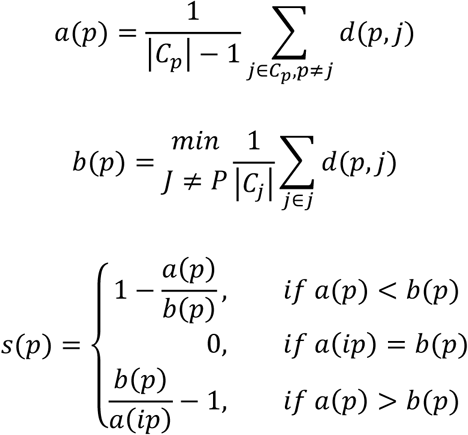

a(i) is the intra-cluster distance. b(i) is the nearest-cluster distance. C is the cluster which patient p belongs to. D represents the Euclidean distance.

#### Endotyping

To generate biologically meaningful and organ system specific endotypes from the computationally defined clusters, we characterized each cluster by its top associated symptoms, inferring the most prominent organ systems. To do this, we computed a cluster-by-symptom matrix using the dot product of the symptom-by-topic matrix and the topic-by-cluster matrix (topic proportions per patient averaged across clusters). The resulting matrix was row-wise normalized (per symptom, across clusters) to highlight the clusters most associated with each symptom, enabling us to categorize (define endotypes) clusters based on their symptom profiles. For each cluster, the top six symptoms with the highest values were selected, corresponding to the darkest squares per column in the heatmap visualization.

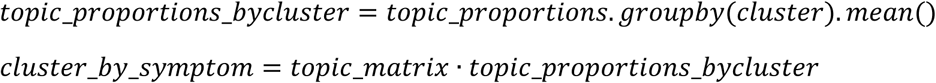

with the dot product of two matrices r and c defined as:

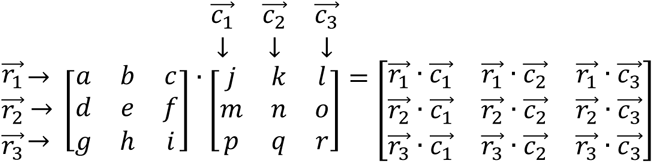

Each element in the result matrix is a dot product of two vectors, r and c. The dot product of two vectors, such as a and b, is defined as:

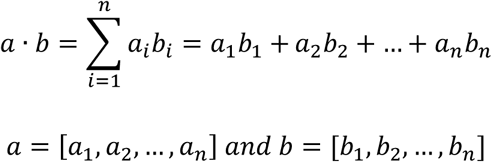

Using ISMMS’s symptom to organ system mapping, we assigned the top three organ systems to each cluster. Symptoms were considered in order of strength, and their corresponding organ systems were collected until a maximum of three distinct organ systems were represented.

To group similar clusters into broader endotypes, we applied a biologically guided agglomerative merging procedure. Referencing the seaborn hierarchical clustermap dendrogram derived from the cluster-by-symptom matrix, the two clusters with the smallest linkage distance (highest similarity) were assessed to determine whether they should be merged based on overlapping organ systems. If at least two out of three organ systems matched or the top organ system matched, the clusters were grouped. This procedure continued iteratively, always merging the next most similar cluster pair based on dendrogram proximity and re-evaluating the last endotype’s top organ systems after each merge. This method led to endotypes comprising one to five clusters, labeled according to their main organs. The data-driven structure uncovered by hierarchical clustering is preserved using this approach, which simultaneously incorporates a domain-specific threshold to ensure organ system interpretability.

There are two possible hierarchical dendrograms, row-normalized and column-normalized. As the endotyping pipeline relies on the distance between clusters, this choice could affect the endotype assignments. Row-normalized results groups clusters based on the most prevalent symptom. Column-normalized results group clusters based on the top contributing symptom for each cluster. This normalization does not affect the top symptoms and, therefore, does not affect the top organ systems. It could affect the order the clusters are potentially merged but the relationships between clusters with the shortest distance remain unchanged. This means only branches with larger distances are flipped, which are unlikely to have similar organ systems anyways.

#### Severity analysis

Severity was defined using the cluster-normalized cluster-by-symptom matrix. This was calculated by subtracting the minimum for each symptom across clusters, then dividing by the difference of the maximum and the minimum. The severity score for each cluster is the sum across symptoms within the normalized matrix. Clusters were ranked by severity score, and a scatter plot was used to visualize the distribution. Based on breaks in the distribution, clusters were grouped into three severity categories: mild, moderate, and severe. This method provides a numerical definition and rank of each cluster’s symptom severity. These groups were used for further statistical analysis comparing assay results and EQ-5D scores. This process was performed for each site (UCSF, ISMMS+Emory, Cardiff) and was used to guide the meta-analysis severity grouping.

These groupings were used to compare the serum and MENSA IgG antibody titer results between severity groups for the Emory cohort (n = 60). Linear regression analysis and Spearman’s correlation were calculated to identify associations between ML-predicted severity scores and mean IgG titer values. One-way ANOVA p-values were reported comparing the mild, moderate, and severe groups (n = 35, 23, and 2, respectively; **Supplementary Figure 3**).

UCSF’s severity groupings were used to compare EQ-5D scores between the two survey timepoints to assess whether patients showed improvement (recovery), no change, or decline (no recovery). EQ-5D scores were chosen for this analysis to represent the overall health of each individual. The inclusion/exclusion criteria for this analysis were: (1) patients must have completed at least one survey with a non-missing EQ-5D score before 85 days and at least one after 85 days; and (2) patients must have indicated the presence of at least one symptom before and after 85 days. These patients are classified as having long COVID. Wilcoxon rank sum statistical testing was performed to evaluate differences in EQ-5D score changes across severity groups and p-values were reported and labeled on the boxplots. This analysis was performed across severity groups, cluster assignments, and endotype assignments.

#### Symptoms over Time

To evaluate the frequency of symptoms over time, we plotted the normalized symptom counts for females and males across three time points (0–85 days, 85 days–365 days, and 365 days on). To account for multiple surveys within a given time period, surveys were combined such that each symptom was recorded as present if at least one survey reported the symptom. For example, if patient A has completed 3 surveys before 85 days, one with cough present and two without, the cough count of patient A would be considered as one. Trends were then assigned into three general categories: decreasing (symptoms reported more at infection but less after 1 year), increasing then decreasing (symptoms increasingly reported after infection and then reducing after 1 year), and increasing (symptoms increasingly reported after infection and after 1 year). These symptoms were categorized based on the percent change of the number of patients who reported the symptom. If the difference between the first and the third time point was less than 25%, that symptom was considered decreasing. If the difference between the first and the third time point was more than 50%, that symptom was considered increasing. If the difference did not fall into the decreasing or increasing category, the symptom was considered increasing then decreasing.

#### Star Charts per Cluster and Median

The purpose pf this analysis was to provide a visual understanding of how similar patient profiles (defined as top six symptoms) were within a cluster. The row-normalized cluster-by-symptom matrix was used to define the top six symptoms for each cluster. The patient-by-symptom matrix was defined as the dot product of the topic matrix and topic proportions. Patient-by-top6symptoms matrix was plotted for each cluster as a star chart. Data from female participants are colored red and male participants colored blue. To show how the presence of the top six symptoms for each cluster were unique, the median values within each cluster were plotted against the median across all other clusters. Cluster 1 was an exception, consistently defined as less symptomatic (control or mild).

Star charts can be coupled with comorbidities proportions for each cluster to suggest correlations between cluster-defining organ systems and known pre-existing conditions.

#### Pie charts

Pie charts were used to visualize differences in cluster, endotype, and severity composition between two groups of patients. For the longitudinal analysis (UCSF only), 162 patients were divided into recovered and non-recovered groups. Recovered was defined as having an increasing EQ-5D score between the first and last surveys, and non-recovered was defined as having a decreasing EQ-5D score. Comparing cluster percentages between recovered and non-recovered groups allowed us to categorize clusters as more or less recovered, providing evidence of predicting changes in symptom severity.

For the meta-analysis, all pre-clustered patients across the four sites (n = 1661) were combined into a large cluster-by-symptom matrix and a cluster-by-organ matrix. This approach was used to redefine the severity groups with the new severity groups containing 398 mild, 1167 moderate, and 96 severe patients. Looking at the pie charts of severity composition for the female and male groups, females made up a majority of moderate and severe cases.

## 5. Discussions

Existing studies have examined symptoms associated with long COVID [Thaweethai 2023, Geng 2024] using a much larger cohort than our study. Relatively consistent long COVID symptom signatures are found in all these studies including ours, including fatigue, brain fog, dizziness, gastrointestinal symptoms, palpitations, change in smell or taste, thirst, chronic cough, chest pain, shortness of breath. Our study furthers the endotyping analysis using ML. In addition to identification of symptom clusters, we associate the symptoms with organ systems, identify severity groups, and compare these patient groups across sites. Our approach demonstrates that all symptoms, regardless of being acute or long COVID, can be modeled together after infection with SARS-CoV-2 for endotyping and severity analysis. In the longitudinal cohort from UCSF, we identified three categories of symptom trajectories: acute resolving, persistent but attenuated, and progressive. Using day 85 as a threshold, we found that some symptom clusters and endotypes improved after three months, while others worsened over time. These findings suggest that the transition from acute to chronic illness is a dynamic continuum rather than discrete phenomena and demonstrate that comprehensive modeling of longitudinal symptom patterns can define clinically meaningful milestones and identify time-sensitive biomarkers for predicting recovery, chronicity, and severity.

Another key challenge in modeling long COVID symptoms is quantifying overall disease severity in a way that enables patient stratification and integration with molecular assay data for biomarker discovery. Traditional health-status measures, such as EQ-5D, lack mapping to specific organ systems and therefore provide limited utility for mechanistic stratification. In the Cardiff case-control cohort, for example, EQ-5D scores failed to distinguish symptom clusters among long COVID cases (**Supplementary Figure 1**). By contrast, our topic modeling-based approach generates data-driven, symptom-derived severity scores that capture clinically meaningful variation. In the ISMMS+Emory cohort, these scores correlated with both EQ-5D (negatively) and MENSA assay data (positively), the latter quantifies SARS-CoV-2-specific antibodies secreted by circulating plasmablasts and provides a sensitive measure of an active immune response. This framework for deriving symptom-based severity scores is broadly applicable and may be adapted to characterize disease severity in other infectious or post-acute-infectious conditions.

An important next step is to identify group-specific biomarkers by linking symptom clusters, endotypes, and severity groups with experimental metadata and assay results. In the Cardiff case-control cohort, we observed notable correlations between severity scores and neutralization capacity against the SARS-CoV-2 Delta variant and a seasonal betacoronavirus, OC43. Interestingly, severity scores did not correlate with neutralization of the ancestral SARS-CoV-2 Wuhan strain, despite lower Wuhan titers in long COVID cases compared to healthy convalescent controls. This discrepancy suggests that variant-specific immune responses rather than overall antibody abundance may be more relevant to the pathogenesis of long COVID. This is reflective of what we found with the Emory study where plasmablasts antibodies may provide a more accurate readout of recent infection compared to pre-existing antibodies. The finding that OC43 neutralizing titers in long COVID cases are higher than those in the control group suggests immunological imprinting established by seasonal betacoronaviruses influences disease outcome. Whether or not pre-existing antibody responses to seasonal coronavirus increases odds of severe disease or is protective requires more in-depth work. Past work from other groups have found a similar trend of higher OC43 binding titers are linked with more severe cases of COVID [Guo 2021] and long COVID [Mak 2025]. Concomitantly, there are also studies that associated higher OC43 antibodies with less severe forms of COVID [Dugas 2021, Kaplonek 2021]. To further explore this possibility, we plan to conduct sex-stratified analyses, including assessment of daily hormone levels and symptoms change, to better understand potential sex-specific mechanisms underlying differences in severity.

Clinical trials are often designed to investigate a single organ system, yet the interrelationships among organ systems in long COVID remain poorly understood. Our meta-analysis, leveraging quantitative topic scores, allowed us to organize symptom clusters across cohorts using a dendrogram, revealing organ system trends. Using this approach, we found that cardiovascular symptoms formed a relatively independent group, suggesting that patients with cardiovascular manifestations may warrant separate study. In contrast, gastrointestinal and pulmonary symptoms co-occurred frequently, indicating that patients with these manifestations could be enrolled together in a unified trial. Such insights can inform the design of new clinical trials and enhance subgroup analyses in ongoing studies, enabling more precise patient stratification based on symptom-defined subgroups.

Our results also demonstrated that TM, rather than traditional dimensionality reduction methods such as PCA or UMAP, is better suited to symptom analysis. It, however, is important to recognize that the TM-based clustering approach has technical limitations. For example, Poisson factorization is stochastic and sensitive to initialization. In our experiments, we found that increasing sample size helped stabilize this process, with likelihood curves becoming smoother and less erratic; enrolling more participants is necessary to further stabilize the results. Averaging results across repeated runs further reduced variability. These improvements were incorporated into our implementation, which we have made publicly available on GitHub. In addition, clustering itself remains a challenge, as there is no universally accepted method to determine the optimal number of clusters. To address this issue, we compared multiple clustering algorithms using silhouette scores and demonstrated consistent results across methods and across cohorts (e.g., ISMMS vs. ISMMS+Emory). This consistency supports the robustness of our approach. Future improvements could involve applying community detection algorithms [Smith 2020] to further partition large clusters (such as Cluster 1 in every cohort), potentially distinguishing controls from mild cases at a finer level of resolution.

## 6. Summary

In this study, we developed and applied a topic modeling-based framework to analyze symptom questionnaires from multiple independent cohorts of patients with long COVID. By transforming binary symptom data into patient-topic representations, we effectively reduced dimensionality, identified reproducible symptom clusters, and organized them into endotypes reflecting organ system involvement. This approach enabled us to derive symptom-based severity scores, which stratify patients into mild, moderate, and severe groups for disease classification. Importantly, these severity scores were significantly associated with independent clinical and immunological measures, including EQ-5D health status scores and MENSA antibody titers, providing external validation of their biological relevance.

Across cohorts, three consistent severity levels were observed, along with cohort-specific endotypes, including gastrointestinal, pulmonary, musculoskeletal, cardiovascular, and hormonal signatures. A female-predominant severe cluster with neurological and hormonal features was repeatedly identified, suggesting sex-specific pathways in long COVID. Longitudinal analysis of the LIINC cohort revealed three distinct symptom trajectories: acute resolving, persistent but attenuated, and progressive, demonstrating that the transition from acute infection to chronic illness is a dynamic continuum.

Meta-analysis further organized global symptom clusters into ten endotypes and revealed relationships among organ systems, with cardiovascular symptoms appearing relatively independent, and gastrointestinal and pulmonary symptoms relating closely. Collectively, these findings highlight the value of computational symptom modeling for deciphering the heterogeneity of long COVID. Moreover, our framework not only provides a reproducible way to cluster patients and quantify severity but also lays the foundation for linking clinical phenotypes to molecular biomarkers, informing mechanistic studies, and optimizing the design and stratification of future clinical trials of targeted interventions for distinct subtypes of long COVID.

## Contributors

RHS, YQ, and YZ initialized the project. YQ and YZ designed the method. YQ, MF, and GST coordinated with participating centers for sample and data transfer. BP implemented the method, processed questionnaires data, and produced analysis results and visualization. YQ conducted statistical analysis and created figures. YQ and BP drafted the manuscript. TD, JNM, JDK, SGD and MJP provided UCSF questionnaires. AR and DP provided ISMMS questionnaires. HED, SAJ, KLM, KL, and DAP provided Cardiff questionnaires. VC, NSH, and FEL provided Emory questionnaires and MENSA data. JE and GST provided neutralization assay data from the Cardiff cohort. CLD, RHS, GST, AP, and MBV provided supervisory suggestions and funding support. HED, DAP, CLD, RHS, YZ, GST, NSH, FEL, AR, DP, MBV, TD, and MJP revised the manuscript. All authors reviewed and approved the manuscript.

## Declaration of interests

ADP and MBV report leadership roles at PolyBio Research Foundation. MJP has received consulting fees from Gilead Sciences, AstraZeneca, BioVie, Apellis Pharmaceuticals, BioNTech, and TechImmune, travel support from Invivyd, and research support from Aerium Therapeutics and Shionogi, outside the submitted work. SGD reports consulting for Enanta Pharmaceuticals and Pfizer and reports research support from Aerium Therapeutics outside the submitted work. YQ reports consulting for Moderna. FEL is the founder of MicroB-plex, Inc. and BeaconDx, Inc., and serves on the scientific board of Be Biopharma. She is a recipient of grants from the BMGF and Genentech, Inc., and has served as a consultant for Astra Zeneca and Novartis. NSH is a scientist at MicroB-plex, Inc. and BeaconDx, Inc., Atlanta, GA. FEL is an inventor of the MENSA patent U.S. Patent No. 10,247,729. April 2, 2019. FEL & NSH are inventors of the MENSA long-COVID diagnostic published patent, March 27, 2025.

## Ethics statement

This research used de-identified questionnaires provided by ISMMS, Emory, Cardiff, and UCSF. All subjects provided written informed consent for inclusion in the studies below. The study protocols at each site were approved by the appropriate Institutional Review Board.

The ISMMS study was approved by the Mount Sinai Program for the Protection of Human Subjects (Study IDs: 23-01715, 21-01147 and 20-01758)

The Emory study was approved under IRB 58271 and was amended/approved for the COVID samples on 3/30/2020.

The Cardiff study was approved by the Cardiff University School of Medicine Research Ethics Committee (21/55) and by the Health Research Authority and Health and Care Research Wales (20/NW/0240).

The UCSF study was approved by the UCSF Institutional Review Board and the UCSF Radiation Safety Committee (ClinicalTirals.gov numbers: NCT04362150 and NCT04815096).

## Data Availability

All data produced in the present study are available upon reasonable request to the authors.

## Acknowledgements

We express our sincere gratitude to all participants, healthcare personnel, study coordinators, administrators, and laboratory managers involved in this work. We thank Catalina Haas Garcia for useful discussions. This work was supported by the PolyBio Research Foundation and the Steven & Alexandra Cohen Foundation, as well as by the U.S. National Institutes of Health (NIH) under grants K23AI157875, R01AI141003, 1R01NS136197, R01AI184931, P01AI168347, U01AI187057, P01AI125180-05S1, R01AI172254, P01AI078907, U01AI045969, U19AI109962, U54CA260563, T32HL116271, 5T32AI074492, and NIH RECOVER 1OT2HL156812-01.

DAP was supported by the UK National Institute for Health and Care Research (COV-LT2-0041) and by the PolyBio Research Foundation (Balvi B43).

This research was also supported in part by the Intramural Research Program of the NIH. The contributions of the NIH author(s) are considered works of the United States Government. The findings and conclusions presented in this paper are those of the author(s) and do not necessarily represent the views of the NIH or the U.S. Department of Health and Human Services.

---

**Supplementary Figure 1.**
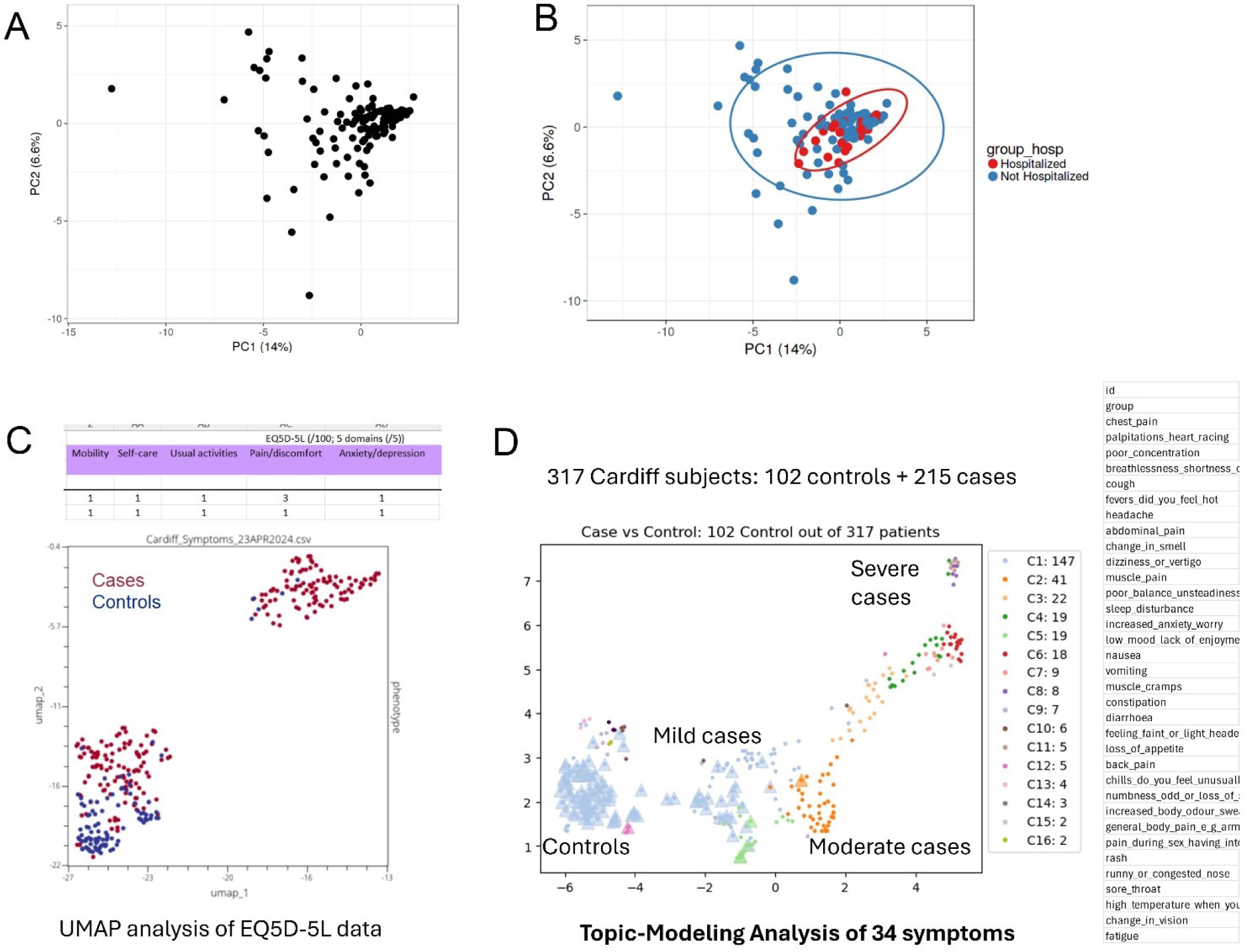
Limitations of PCA, UMAP, and EQ-5D scores for identifying questionnaire-based symptom clusters or predicting severity without topic modeling. **(A)** Principal component analysis (PCA) of 144 UCSF participants across 32 symptoms fails to reveal distinct clusters. **(B)** PCA also cannot distinguish hospitalized from non-hospitalized patients. **(C)** EQ-5D-5L scores from the Cardiff identify only two patient groups via UMAP, with overlap between cases and controls. **(D)** In contrast, applying UMAP to the topic modeling-transformed patient-by-topic matrix separates cases from controls in the Cardiff cohort and further identifies multiple symptom clusters among participants with long COVID.

**Supplementary Figure 2.**
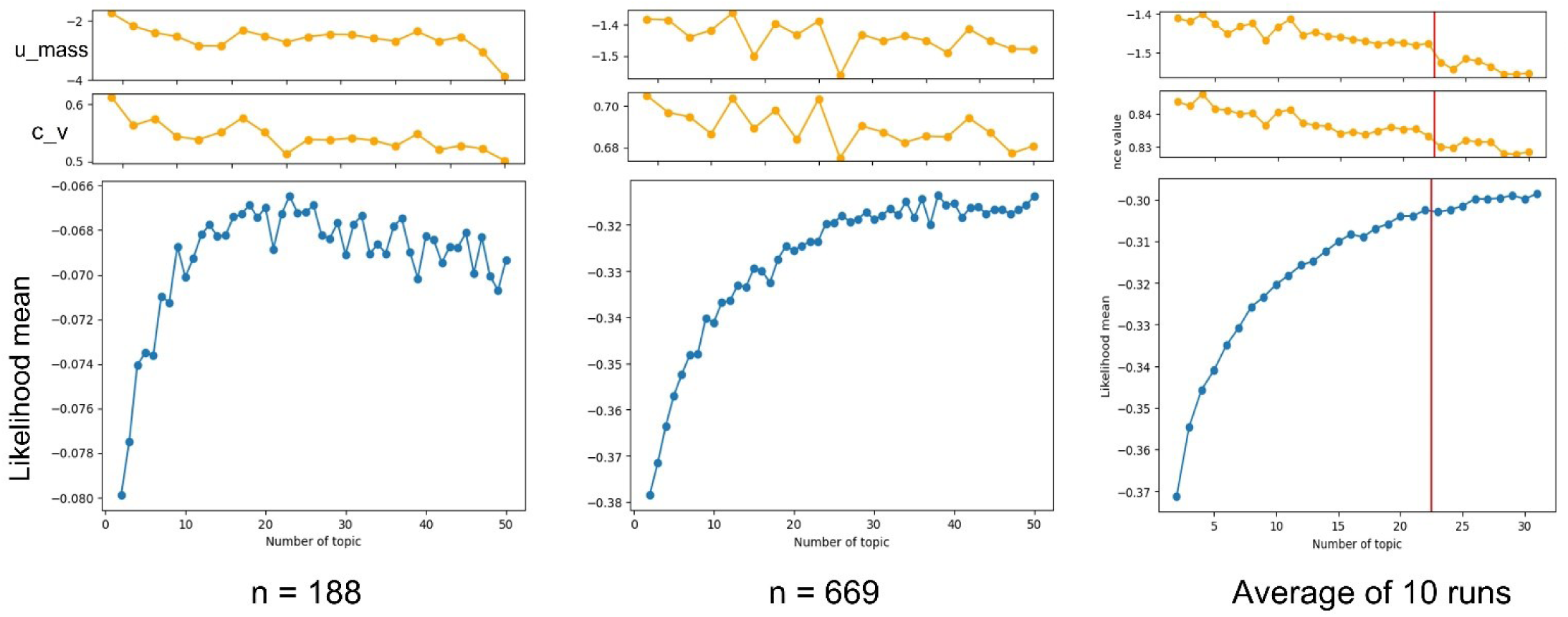
Balancing topic coherence and data likelihood for selecting the optimal number of topics in topic modeling. *U_mass* and *c_v* (yellow lines, upper region of Y-axis) are coherence metrics that quantify intra-cluster similarity, while likelihood (blue line, lower region of Y-axis) reflects how well the data are partitioned. Higher values for both indicate better feature clustering. As the number of topics (X-axis) increases, likelihood generally rises, whereas coherence tends to decline. The required number of topics typically scales with data size. Using the UCSF data as an example: when *n* = 188, likelihood plateaus beyond ∼20 topics; when *n* = 669, likelihood continues improving up to ∼40 topics. By averaging the results over ten repeated runs, smoothing the curves, and evaluating the combined coherence-likelihood score, we determined the optimal topic number to be 22.

**Supplementary Figure 3.**
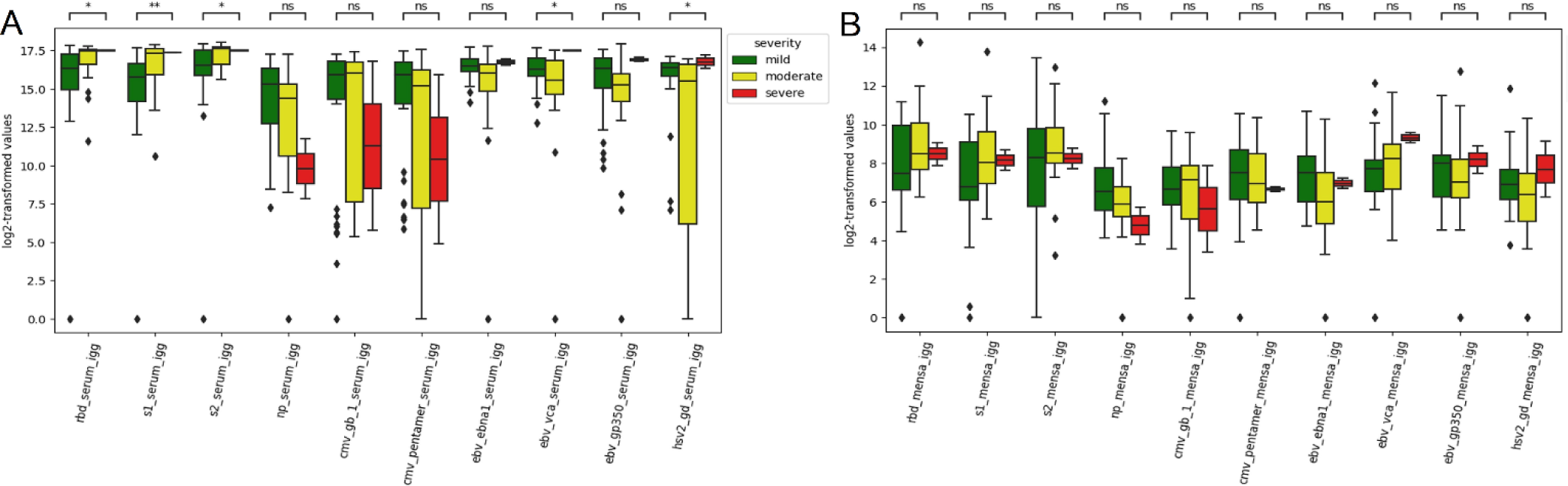
Serum and MENSA antibody assay results stratified by severity across ten experiments. The X-axis shows three severity groups (mild: green; moderate: yellow; severe: red), and the Y-axis shows log₂-transformed IgG titers. **(A)** Serum assays and **(B)** MENSA assays are displayed as box plots, with the median, standard deviation, and outliers (diamonds) indicated. IgG titers generally increase with higher severity. Differences among severity groups were assessed using a non-parametric one-way ANOVA (Kruskal-Wallis test). In the serum data, significant differences were observed in responses to the RBD, S1, and S2 regions of SARS-CoV-2, as well as in responses to EBV and HSV-2.

## Notes

### Competing Interest Statement

ADP and MBV reports leadership roles at PolyBio Research Foundation. MJP has received consulting fees from Gilead Sciences, AstraZeneca, BioVie, Apellis Pharmaceuticals, BioNTech, and TechImmune, travel support from Invivyd, and research support from Aerium Therapeutics and Shionogi, outside the submitted work. SGD reports consulting for Enanta Pharmaceuticals and Pfizer and reports research support from Aerium Therapeutics outside the submitted work. YQ reports consulting for Moderna. FEL is the founder of MicroB-plex, Inc. and BeaconDx, Inc., and serves on the scientific board of Be Biopharma. She is a recipient of grants from the BMGF and Genentech, Inc., and has served as a consultant for Astra Zeneca and Novartis. NSH is a scientist at MicroB-plex, Inc. and BeaconDx, Inc., Atlanta, GA. FEL is an inventor of the MENSA patent U.S. Patent No. 10,247,729. April 2, 2019. FEL & NSH are inventors of the MENSA long-COVID diagnostic published patent, March 27, 2025.

### Author Declarations

This research used de-identified questionnaires provided by ISMMS, Emory, Cardiff, and UCSF. All subjects provided written informed consent for inclusion in the studies below. The study protocols at each site were approved by the appropriate Institutional Review Board. The ISMMS study was approved by the Mount Sinai Program for the Protection of Human Subjects (Study IDs: 23-01715, 21-01147 and 20-01758) The Emory study was approved under IRB 58271 and was amended/approved for the COVID samples on 3/30/2020. The Cardiff study was approved by the Cardiff University School of Medicine Research Ethics Committee (21/55) and by the Health Research Authority and Health and Care Research Wales (20/NW/0240). The UCSF study was approved by the UCSF Institutional Review Board and the UCSF Radiation Safety Committee (ClinicalTirals.gov numbers: NCT04362150 and NCT04815096).

### Summary of Updates

1.Authors (except for the first three and last two authors) are now listed alphabetically. 2.New information added to figures: Figure 1 now includes schematic illustrations of clusters, endotypes, and severity groups. A new Figure 6 has been added to visualize longitudinal trajectories of symptom clusters. Figure 5 has been updated accordingly, with corresponding updates to figure legends and text. 3.Risk factor analysis. We added a risk factor analysis showing that individuals with non-mild symptoms (as identified by the machine learning approach) during the acute phase had a higher risk of developing moderate or severe long COVID compared with individuals with mild acute symptoms. 4.Minor corrections. We corrected numerous spelling errors and mislabeled items throughout the text, figures, and figure legends. Acknowledgement is also updated with most recent information.

## References

Ahmad I, Amelio A, Merla A, Scozzari F. A survey on the role of artificial intelligence in managing Long COVID. Front Artif Intell. 2024 Jan 11;6:1292466. doi: 10.3389/frai.2023.1292466. PMID: 38274052; PMCID: PMC10808521.

Baillie K, Davies HE, Keat SBK, Ladell K, Miners KL, Jones SA, Mellou E, Toonen EJM, Price DA, Morgan BP, Zelek WM. Complement dysregulation is a prevalent and therapeutically amenable feature of long COVID. Med. 2024 Mar 8;5(3):239–253.e5. doi: 10.1016/j.medj.2024.01.011. Epub 2024 Feb 15. PMID: 38359836.

Bowe B, Xie Y, Al-Aly Z. Postacute sequelae of COVID-19 at 2 years. Nat Med. 2023 Sep;29(9):2347–2357. doi: 10.1038/s41591-023-02521-2. Epub 2023 Aug 21. PMID: 37605079; PMCID: PMC10504070.

Chippa V, Aleem A, Anjum F. Postacute Coronavirus (COVID-19) Syndrome. 2024 Mar 19. In: StatPearls [Internet]. Treasure Island (FL): StatPearls Publishing; 2025 Jan–. PMID: 34033370.

Constantinescu-Bercu A, Lobiuc A, Căliman-Sturdza OA, Oiţă RC, Iavorschi M, Pavăl NE, Șoldănescu I, Dimian M, Covasa M. Long COVID: Molecular Mechanisms and Detection Techniques. Int J Mol Sci. 2023 Dec 28;25(1):408. doi: 10.3390/ijms25010408. PMID: 38203577; PMCID: PMC10778767.

Davis HE, McCorkell L, Vogel JM, Topol EJ. Long COVID: major findings, mechanisms and recommendations. Nat Rev Microbiol. 2023 Mar;21(3):133–146. doi: 10.1038/s41579-022-00846-2. Epub 2023 Jan 13. Erratum in: Nat Rev Microbiol. 2023 Jun;21(6):408. doi: 10.1038/s41579-023-00896-0. PMID: 36639608; PMCID: PMC9839201.

Devlin N, Parkin D, Janssen B. Methods for Analysing and Reporting EQ-5D Data [Internet]. Cham (CH): Springer; 2020. PMID: 33347096.

Diaz-Papkovich A, Anderson-Trocmé L, Gravel S. A review of UMAP in population genetics. J Hum Genet. 2021 Jan;66(1):85–91. doi: 10.1038/s10038-020-00851-4. Epub 2020 Oct 14. PMID: 33057159; PMCID: PMC7728596.

Dugas M, Grote-Westrick T, Vollenberg R, Lorentzen E, Brix T, Schmidt H, Tepasse PR, Kühn J. Less severe course of COVID-19 is associated with elevated levels of antibodies against seasonal human coronaviruses OC43 and HKU1 (HCoV OC43, HCoV HKU1). Int J Infect Dis. 2021 Apr;105:304–306. doi: 10.1016/j.ijid.2021.02.085. Epub 2021 Feb 23. PMID: 33636357; PMCID: PMC7901274.

Feng Y, Parkin D, Devlin NJ. Assessing the performance of the EQ-VAS in the NHS PROMs programme. Qual Life Res. 2014 Apr;23(3):977–89. doi: 10.1007/s11136-013-0537-z. Epub 2013 Oct 1. PMID: 24081873; PMCID: PMC4287662.

Fresi E, Pagani E, Pezzetti F, Montomoli C, Monti C, Betti M, De Silvestri A, Sagliocco O, Zuccaro V, Bruno R, Klersy C. Long COVID’s Hidden Complexity: Machine Learning Reveals Why Personalized Care Remains Essential. J Clin Med. 2025 May 23;14(11):3670. doi: 10.3390/jcm14113670. PMID: 40507431; PMCID: PMC12155299.

Frontera JA, Thorpe LE, Simon NM, de Havenon A, Yaghi S, Sabadia SB, Yang D, Lewis A, Melmed K, Balcer LJ, Wisniewski T, Galetta SL. Post-acute sequelae of COVID-19 symptom phenotypes and therapeutic strategies: A prospective, observational study. PLoS One. 2022 Sep 29;17(9):e0275274. doi: 10.1371/journal.pone.0275274. PMID: 36174032; PMCID: PMC9521913.

Gao Y, Cai C, Adamo S, Biteus E, Kamal H, Dager L, Miners KL, Llewellyn-Lacey S, Ladell K, Amratia PS, Bentley K, Kollnberger S, Wu J, Akhirunnesa M, Jones SA, Julin P, Lidman C, Stanton RJ, Goepfert PA, Peluso MJ, Deeks SG, Davies HE, Aleman S, Buggert M, Price DA. Identification of soluble biomarkers that associate with distinct manifestations of long COVID. Nat Immunol. 2025 May;26(5):692–705. doi: 10.1038/s41590-025-02135-5. Epub 2025 Apr 30. PMID: 40307449; PMCID: PMC12043503.

Gan, Z., Chen, C., Henao, R., Carlson, D., Carin, L. (2015). Scalable Deep Poisson Factor Analysis for Topic Modeling. Proceedings of Machine Learning Research. PMLR 37:1823–1832, 2015.

Gáspár Z, Szabó BG, Ceglédi A, Lakatos B. Human herpesvirus reactivation and its potential role in the pathogenesis of post-acute sequelae of SARS-CoV-2 infection. Geroscience. 2025 Feb;47(1):167–187. doi: 10.1007/s11357-024-01323-9. Epub 2024 Aug 29. PMID: 39207648; PMCID: PMC11872864.

Geng LN, Erlandson KM, Hornig M, Letts R, Selvaggi C, Ashktorab H, Atieh O, Bartram L, Brim H, Brosnahan SB, Brown J, Castro M, Charney A, Chen P, Deeks SG, Erdmann N, Flaherman VJ, Ghamloush MA, Goepfert P, Goldman JD, Han JE, Hess R, Hirshberg E, Hoover SE, Katz SD, Kelly JD, Klein JD, Krishnan JA, Lee-Iannotti J, Levitan EB, Marconi VC, Metz TD, Modes ME, Nikolich JŽ, Novak RM, Ofotokun I, Okumura MJ, Parthasarathy S, Patterson TF, Peluso MJ, Poppas A, Quintero Cardona O, Scott J, Shellito J, Sherif ZA, Singer NG, Taylor BS, Thaweethai T, Verduzco-Gutierrez M, Wisnivesky J, McComsey GA, Horwitz LI, Foulkes AS; RECOVER Consortium. 2024 Update of the RECOVER-Adult Long COVID Research Index. JAMA. 2025 Feb 25;333(8):694–700. doi: 10.1001/jama.2024.24184. PMID: 39693079; PMCID: PMC11862971.

Guo L, Wang Y, Kang L, Hu Y, Wang L, Zhong J, Chen H, Ren L, Gu X, Wang G, Wang C, Dong X, Wu C, Han L, Wang Y, Fan G, Zou X, Li H, Xu J, Jin Q, Cao B, Wang J. Cross-reactive antibody against human coronavirus OC43 spike protein correlates with disease severity in COVID-19 patients: a retrospective study. Emerg Microbes Infect. 2021 Dec;10(1):664–676. doi: 10.1080/22221751.2021.1905488. PMID: 33734013; PMCID: PMC8023607.

Haddad NS, Morrison-Porter A, Quehl H, Capric V, Lamothe PA, Anam F, Runnstrom MC, Truong AD, Dixit AN, Woodruff MC, Chen A, Park J, Nguyen DC, Hentenaar I, Kim CY, Kyu S, Stewart B, Wagman E, Geoffroy H, Sanz D, Cashman KS, Ramonell RP, Cabrera-Mora M, Alter DN, Roback JD, Horwath MC, O’Keefe JB, Dretler AW, Gripaldo R, Yeligar SM, Natoli T, Betin V, Patel R, Vela K, Hernandez MR, Usman S, Varghese J, Jalal A, Lee S, Le SN, Amoss RT, Daiss JL, Sanz I, Lee FE. MENSA, a Media Enriched with Newly Synthesized Antibodies, to Identify SARS-CoV-2 Persistence and Latent Viral Reactivation in Long-COVID. medRxiv [Preprint]. 2024 Jul 7:2024.07.05.24310017. doi: 10.1101/2024.07.05.24310017. PMID: 39006446; PMCID: PMC11245097.

Hill EL, Mehta HB, Sharma S, Mane K, Singh SK, Xie C, Cathey E, Loomba J, Russell S, Spratt H, DeWitt PE, Ammar N, Madlock-Brown C, Brown D, McMurry JA, Chute CG, Haendel MA, Moffitt R, Pfaff ER, Bennett TD; N3C Consortium; and the RECOVER Consortium. Risk factors associated with post-acute sequelae of SARS-CoV-2: an N3C and NIH RECOVER study. BMC Public Health. 2023 Oct 25;23(1):2103. doi: 10.1186/s12889-023-16916-w. PMID: 37880596; PMCID: PMC10601201.

Kaplonek P, Wang C, Bartsch Y, Fischinger S, Gorman MJ, Bowman K, Kang J, Dayal D, Martin P, Nowak RP, Villani AC, Hsieh CL, Charland NC, Gonye ALK, Gushterova I, Khanna HK, LaSalle TJ, Lavin-Parsons KM, Lilley BM, Lodenstein CL, Manakongtreecheep K, Margolin JD, McKaig BN, Rojas-Lopez M, Russo BC, Sharma N, Tantivit J, Thomas MF, Sade-Feldman M, Feldman J, Julg B, Nilles EJ, Musk ER, Menon AS, Fischer ES, McLellan JS, Schmidt A, Goldberg MB, Filbin MR, Hacohen N, Lauffenburger DA, Alter G. Early cross-coronavirus reactive signatures of humoral immunity against COVID-19. Sci Immunol. 2021 Oct 15;6(64):eabj2901. doi: 10.1126/sciimmunol.abj2901. Epub 2021 Oct 15. PMID: 34652962; PMCID: PMC8943686.

Lopez-Leon S, Wegman-Ostrosky T, Perelman C, Sepulveda R, Rebolledo PA, Cuapio A, Villapol S. More than 50 long-term effects of COVID-19: a systematic review and meta-analysis. Sci Rep. 2021 Aug 9;11(1):16144. doi: 10.1038/s41598-021-95565-8. PMID: 34373540; PMCID: PMC8352980.

Mak WA, Wapperom D, Redel AL, Koeleman JGM, Smit PW, Lam-Tse WK, van der Poll T, Chen HJ, den Dunnen J, Braunstahl GJ, Ong DSY. Seasonal Coronavirus-Induced Immunological Imprinting and Previous Herpesvirus Infections in Patients With Long COVID. J Med Virol. 2025 Sep;97(9):e70582. doi: 10.1002/jmv.70582. PMID: 40884088; PMCID: PMC12397718.

National Academies of Sciences, Engineering, and Medicine; Health and Medicine Division; Board on Global Health; Board on Health Sciences Policy; Committee on Examining the Working Definition for Long COVID. A Long COVID Definition: A Chronic, Systemic Disease State with Profound Consequences. Goldowitz I, Worku T, Brown L, Fineberg HV, editors. Washington (DC): National Academies Press (US); 2024 Jul 9. PMID: 39110819.

O’Neil ST, Madlock-Brown C, Wilkins KJ, McGrath BM, Davis HE, Assaf GS, Wei H, Zareie P, French ET, Loomba J, McMurry JA, Zhou A, Chute CG, Moffitt RA, Pfaff ER, Yoo YJ, Leese P, Chew RF, Lieberman M, Haendel MA; N3C and RECOVER Consortia. Finding Long-COVID: temporal topic modeling of electronic health records from the N3C and RECOVER programs. NPJ Digit Med. 2024 Oct 21;7(1):296. doi: 10.1038/s41746-024-01286-3. PMID: 39433942; PMCID: PMC11494196.

Parotto M, Gyöngyösi M, Howe K, Myatra SN, Ranzani O, Shankar-Hari M, Herridge MS. Post-acute sequelae of COVID-19: understanding and addressing the burden of multisystem manifestations. Lancet Respir Med. 2023 Aug;11(8):739–754. doi: 10.1016/S2213-2600(23)00239-4. Epub 2023 Jul 17. PMID: 37475125.

Peluso MJ, Kelly JD, Lu S, Goldberg SA, Davidson MC, Mathur S, Durstenfeld MS, Spinelli MA, Hoh R, Tai V, Fehrman EA, Torres L, Hernandez Y, Williams MC, Arreguin MI, Ngo LH, Deswal M, Munter SE, Martinez EO, Anglin KA, Romero MD, Tavs J, Rugart PR, Chen JY, Sans HM, Murray VW, Ellis PK, Donohue KC, Massachi JA, Weiss JO, Mehdi I, Pineda-Ramirez J, Tang AF, Wenger MA, Assenzio MT, Yuan Y, Krone MR, Rutishauser RL, Rodriguez-Barraquer I, Greenhouse B, Sauceda JA, Gandhi M, Scheffler AW, Hsue PY, Henrich TJ, Deeks SG, Martin JN. Persistence, Magnitude, and Patterns of Postacute Symptoms and Quality of Life Following Onset of SARS-CoV-2 Infection: Cohort Description and Approaches for Measurement. Open Forum Infect Dis. 2021 Dec 21;9(2):ofab640. doi: 10.1093/ofid/ofab640. PMID: 35106317; PMCID: PMC8755302.

Peluso MJ, Ryder D, Flavell RR, Wang Y, Levi J, LaFranchi BH, Deveau TM, Buck AM, Munter SE, Asare KA, Aslam M, Koch W, Szabo G, Hoh R, Deswal M, Rodriguez AE, Buitrago M, Tai V, Shrestha U, Lu S, Goldberg SA, Dalhuisen T, Vasquez JJ, Durstenfeld MS, Hsue PY, Kelly JD, Kumar N, Martin JN, Gambhir A, Somsouk M, Seo Y, Deeks SG, Laszik ZG, VanBrocklin HF, Henrich TJ. Tissue-based T cell activation and viral RNA persist for up to 2 years after SARS-CoV-2 infection. Sci Transl Med. 2024 Jul 3;16(754):eadk3295. doi: 10.1126/scitranslmed.adk3295. Epub 2024 Jul 3. PMID: 38959327; PMCID: PMC11337933.

Peluso MJ, Deeks SG. Mechanisms of long COVID and the path toward therapeutics. Cell. 2024 Oct 3;187(20):5500–5529. doi: 10.1016/j.cell.2024.07.054. Epub 2024 Sep 25. PMID: 39326415; PMCID: PMC11455603.

Prakash S, Karan S, Lekbach Y, Tifrea DF, Figueroa CJ, Ulmer JB, Young JF, Glenn G, Gil D, Jones TM, Redfield RR, BenMohamed L. Insights into Persistent SARS-CoV-2 Reservoirs in Chronic Long COVID. Viruses. 2025 Sep 27;17(10):1310. doi: 10.3390/v17101310. PMID: 41157582; PMCID: PMC12568064.

Proal AD, VanElzakker MB, Aleman S, Bach K, Boribong BP, Buggert M, Cherry S, Chertow DS, Davies HE, Dupont CL, Deeks SG, Eimer W, Ely EW, Fasano A, Freire M, Geng LN, Griffin DE, Henrich TJ, Iwasaki A, Izquierdo-Garcia D, Locci M, Mehandru S, Painter MM, Peluso MJ, Pretorius E, Price DA, Putrino D, Scheuermann RH, Tan GS, Tanzi RE, VanBrocklin HF, Yonker LM, Wherry EJ. SARS-CoV-2 reservoir in post-acute sequelae of COVID-19 (PASC). Nat Immunol. 2023 Oct;24(10):1616–1627. doi: 10.1038/s41590-023-01601-2. Epub 2023 Sep 4. Erratum in: Nat Immunol. 2023 Oct;24(10):1778. doi: 10.1038/s41590-023-01646-3. PMID: 37667052.

Proal AD, Aleman S, Bomsel M, Brodin P, Buggert M, Cherry S, Chertow DS, Davies HE, Dupont CL, Deeks SG, Ely EW, Fasano A, Freire M, Geng LN, Griffin DE, Henrich TJ, Hewitt SM, Iwasaki A, Krumholz HM, Locci M, Marconi VC, Mehandru S, Muller-Trutwin M, Painter MM, Pretorius E, Price DA, Putrino D, Qian Y, Roan NR, Salmon D, Tan GS, VanElzakker MB, Wherry EJ, Van Weyenbergh J, Yonker LM, Peluso MJ. Targeting the SARS-CoV-2 reservoir in long COVID. Lancet Infect Dis. 2025 May;25(5):e294–e306. doi: 10.1016/S1473-3099(24)00769-2. Epub 2025 Feb 10. PMID: 39947217; PMCID: PMC12151306.

Röder, M., Both, A, Hinneburg A. Exploring the Space of Topic Coherence Measures. Proceedings of the Eighth ACM International Conference on Web Search and Data Mining (2015), pp. 399–408.

Rousseeuw, Peter. (1987). Silhouettes: A graphical aid to the interpretation and validation of cluster analysis. Journal of Computational and Applied Mathematics. Volume 20, pp. 53–65. 10.1016/0377-0427(87)90125-7.

Smith NR, Zivich PN, Frerichs LM, Moody J, Aiello AE. A Guide for Choosing Community Detection Algorithms in Social Network Studies: The Question Alignment Approach. Am J Prev Med. 2020 Oct;59(4):597–605. doi: 10.1016/j.amepre.2020.04.015. PMID: 32951683; PMCID: PMC7508227.

Soriano JB, Murthy S, Marshall JC, Relan P, Diaz JV; WHO Clinical Case Definition Working Group on Post-COVID-19 Condition. A clinical case definition of post-COVID-19 condition by a Delphi consensus. Lancet Infect Dis. 2022 Apr;22(4):e102–e107. doi: 10.1016/S1473-3099(21)00703-9. Epub 2021 Dec 21. PMID: 34951953; PMCID: PMC8691845.

Swank Z, Borberg E, Chen Y, Senussi Y, Chalise S, Manickas-Hill Z, Yu XG, Li JZ, Alter G, Henrich TJ, Kelly JD, Hoh R, Goldberg SA, Deeks SG, Martin JN, Peluso MJ, Talla A, Li X, Skene P, Bumol TF, Torgerson TR, Czartoski JL, McElrath MJ, Karlson EW, Walt DR; RECOVER consortium authors. Measurement of circulating viral antigens post-SARS-CoV-2 infection in a multicohort study. Clin Microbiol Infect. 2024 Dec;30(12):1599–1605. doi: 10.1016/j.cmi.2024.09.001. Epub 2024 Oct 9. PMID: 39389851; PMCID: PMC11578795.

Thaweethai T, Jolley SE, Karlson EW, Levitan EB, Levy B, McComsey GA, McCorkell L, Nadkarni GN, Parthasarathy S, Singh U, Walker TA, Selvaggi CA, Shinnick DJ, Schulte CCM, Atchley-Challenner R, Alba GA, Alicic R, Altman N, Anglin K, Argueta U, Ashktorab H, Baslet G, Bassett IV, Bateman L, Bedi B, Bhattacharyya S, Bind MA, Blomkalns AL, Bonilla H, Brim H, Bush PA, Castro M, Chan J, Charney AW, Chen P, Chibnik LB, Chu HY, Clifton RG, Costantine MM, Cribbs SK, Davila Nieves SI, Deeks SG, Duven A, Emery IF, Erdmann N, Erlandson KM, Ernst KC, Farah-Abraham R, Farner CE, Feuerriegel EM, Fleurimont J, Fonseca V, Franko N, Gainer V, Gander JC, Gardner EM, Geng LN, Gibson KS, Go M, Goldman JD, Grebe H, Greenway FL, Habli M, Hafner J, Han JE, Hanson KA, Heath J, Hernandez C, Hess R, Hodder SL, Hoffman MK, Hoover SE, Huang B, Hughes BL, Jagannathan P, John J, Jordan MR, Katz SD, Kaufman ES, Kelly JD, Kelly SW, Kemp MM, Kirwan JP, Klein JD, Knox KS, Krishnan JA, Kumar A, Laiyemo AO, Lambert AA, Lanca M, Lee-Iannotti JK, Logarbo BP, Longo MT, Luciano CA, Lutrick K, Maley JH, Mallett G, Marathe JG, Marconi V, Marshall GD, Martin CF, Matusov Y, Mehari A, Mendez-Figueroa H, Mermelstein R, Metz TD, Morse R, Mosier J, Mouchati C, Mullington J, Murphy SN, Neuman RB, Nikolich JZ, Ofotokun I, Ojemakinde E, Palatnik A, Palomares K, Parimon T, Parry S, Patterson JE, Patterson TF, Patzer RE, Peluso MJ, Pemu P, Pettker CM, Plunkett BA, Pogreba-Brown K, Poppas A, Quigley JG, Reddy U, Reece R, Reeder H, Reeves WB, Reiman EM, Rischard F, Rosand J, Rouse DJ, Ruff A, Saade G, Sandoval GJ, Santana JL, Schlater SM, Sciurba FC, Shepherd F, Sherif ZA, Simhan H, Singer NG, Skupski DW, Sowles A, Sparks JA, Sukhera FI, Taylor BS, Teunis L, Thomas RJ, Thorp JM, Thuluvath P, Ticotsky A, Tita AT, Tuttle KR, Urdaneta AE, Valdivieso D, VanWagoner TM, Vasey A, Verduzco-Gutierrez M, Wallace ZS, Ward HD, Warren DE, Weiner SJ, Welch S, Whiteheart SW, Wiley Z, Wisnivesky JP, Yee LM, Zisis S, Horwitz LI, Foulkes AS; RECOVER Consortium. Development of a Definition of Postacute Sequelae of SARS-CoV-2 Infection. JAMA. 2023 Jun 13;329(22):1934–1946. doi: 10.1001/jama.2023.8823. PMID: 37278994; PMCID: PMC10214179.

Zhang H, Zang C, Xu Z, Zhang Y, Xu J, Bian J, Morozyuk D, Khullar D, Zhang Y, Nordvig AS, Schenck EJ, Shenkman EA, Rothman RL, Block JP, Lyman K, Weiner MG, Carton TW, Wang F, Kaushal R. Data-driven identification of post-acute SARS-CoV-2 infection subphenotypes. Nat Med. 2023 Jan;29(1):226–235. doi: 10.1038/s41591-022-02116-3. Epub 2022 Dec 1. PMID: 36456834; PMCID: PMC9873564.

Zhou, M., Hannah, L., Dunson, D. & Carin, L. Beta-negative binomial process and Poisson factor analysis. Proceedings of the Fifteenth International Conference on Artificial Intelligence and Statistics, PMLR 22:1462–1471 (2012).

